# Comparison of algorithm-based versus single-item diagnostic measures of anxiety and depression disorders in the GLAD and COPING cohorts

**DOI:** 10.1101/2021.01.08.21249434

**Authors:** Molly R. Davies, Joshua E. J. Buckman, Brett N. Adey, Chérie Armour, John R. Bradley, Susannah C. B. Curzons, Katrina A. S. Davis, Kimberley A. Goldsmith, Colette R. Hirsch, Matthew Hotopf, Christopher Hübel, Ian R. Jones, Gursharan Kalsi, Georgina Krebs, Yuhao Lin, Ian Marsh, Monika McAtarsney-Kovacs, Andrew M. McIntosh, Dina Monssen, Alicia J. Peel, Henry C. Rogers, Megan Skelton, Daniel J. Smith, Abigail ter Kuile, Katherine N. Thompson, David Veale, James T. R. Walters, Roland Zahn, Gerome Breen, Thalia C. Eley

**Affiliations:** Institute of Psychiatry, Psychology and Neuroscience, King’s College London, Denmark Hill, Camberwell, London, UK; National Institute for Health Research (NIHR) Biomedical Research Centre, South London and Maudsley Hospital, London, UK; Centre for Outcomes Research and Effectiveness (CORE), Research Department of Clinical, Educational and Health Psychology, University College London, Gower Street, London WC1E 7HB, United Kingdom; iCope – Camden and Islington Psychological Therapies Services, Camden & Islington NHS Foundation Trust, St Pancras Hospital, London, UK; Stress, Trauma & Related Conditions (STARC) research lab, School of Psychology, Queens University Belfast (QUB), Belfast, Northern Ireland, UK; NIHR BioResource, Cambridge University Hospitals, Cambridge Biomedical Campus, Cambridge, UK; South London and Maudsley NHS Foundation Trust, Denmark Hill, Camberwell, London, UK; Department of Medical Epidemiology and Biostatistics, Karolinska Institutet, Stockholm, Sweden; National Centre for Register-based Research, Aarhus Business and Social Sciences, Aarhus University, Aarhus, Denmark; National Centre for Mental Health, Division of Psychiatry and Clinical Neuroscience, Cardiff University, Cardiff, UK; Division of Psychiatry, Centre for Clinical Brain Sciences, University of Edinburgh, Edinurgh, UK; Institute of Health and Wellbeing, University of Glasgow, Glasgow, UK

## Abstract

**Background:** Understanding and improving outcomes for people with anxiety or depression often requires large studies. To increase participation and reduce costs, such research is typically unable to utilise “gold-standard” methods to ascertain diagnoses, instead relying on remote, self-report measures.

**Aims:** To assess the comparability of remote diagnostic methods for anxiety and depression disorders commonly used in research.

**Method:** Participants from the UK-based GLAD and COPING NBR cohorts (*N* = 58,400) completed an online questionnaire between 2018-2020. Responses to detailed symptom reports were compared to DSM-5 criteria to generate algorithm-based diagnoses of major depressive disorder (MDD), generalised anxiety disorder (GAD), specific phobia, social anxiety disorder, panic disorder, and agoraphobia. Participants also self-reported any prior diagnoses from health professionals, termed single-item diagnoses. “Any anxiety” included participants with at least one anxiety disorder. Agreement was assessed by calculating accuracy, Cohen’s kappa, McNemar’s chi-squared, sensitivity, and specificity.

**Results:** Agreement between diagnoses was moderate for MDD, any anxiety, and GAD, but varied by cohort. Agreement was slight to fair for the phobic disorders. Many participants with single-item GAD did not receive an algorithm-based diagnosis. In contrast, algorithm-based diagnoses of the phobic disorders were more common than single-item diagnoses.

**Conclusions:** Agreement for MDD, any anxiety, and GAD was higher for cases in the case-enriched GLAD cohort and for controls in the general population COPING NBR cohort. For anxiety disorders, single-item diagnoses classified most participants as having GAD, whereas algorithm-based diagnoses distributed participants more evenly across the anxiety disorders. Further validation against gold standard measures is required.

## Introduction

Anxiety and depressive disorders are common and debilitating, impacting approximately 30% of the population during their lifetime (1,2), and accounting for 10% of years lived with disability (3). This highlights the importance of understanding disorder-related risk factors and outcomes. In order to undertake research or treatment of these conditions, a vital step is identifying participants with or without the disorder of interest. The “gold standard” for ascertaining disorder diagnoses in psychiatric research is a structured or semi-structured diagnostic interview conducted in person or over the phone by a trained interviewer, such as the Composite International Diagnostic Interview (CIDI) (4) or Structured Clinical Interview for DSM-5 (SCID) (5). However, conducting interviews is time-consuming and costly. Due to the heterogeneous and complex aetiology of anxiety and depression, studies often require extremely large samples to reach sufficient statistical power. This renders diagnostic interviews impractical, and large-scale studies increasingly use online, self-report questionnaires to ascertain anxiety and depressive disorder diagnostic status of participants.

There are two common methods to ascertain a diagnosis when using online questionnaires. *Algorithm-based diagnoses* involve a questionnaire which asks participants to self-report specific symptoms. The questionnaire responses are run through an algorithm and compared to diagnostic criteria, such as the Diagnostic Statistical Manual (DSM-5; (6), to assess whether the participant meets criteria for a diagnosis. This has been referred to as either strictly-defined, detailed, or symptom-based diagnosis (7,8). *Single-item diagnoses* take a contrasting approach and utilise a single question where participants are asked about whether they have received a clinical diagnosis from a health professional for a psychiatric disorder during their lifetime. This is also known as minimal, broad, or light-touch diagnosis (8,9). Both algorithm-based and single-item diagnostic methods are in widespread use in anxiety and depression research; however, it is unclear whether they identify the same individuals.

One field which has focused heavily on ways to ascertain large sample sizes is that of psychiatric genetics. Anxiety and depression are heritable disorders. Heritability refers to the proportion of variance of a trait or disorder that is attributed to genetic factors, and decades of work have shown that approximately 20-30% of the variance of anxiety and depression can be attributed to genetics (10–13). Due to the complex nature of the genetics of anxiety and depression disorders, samples in the hundreds of thousands have been required for adequate statistical power to detect significant associations with genetic variants. Psychiatric genetic studies have therefore used a variety of approaches to determine case and control status, often combining multiple methods in meta-analyses, and have endeavoured to assess whether these measures represent the same constructs by investigating the genetic correlation between them. Genetic correlations indicate whether the same genetic variants are associated with different traits, thus lending evidence about their similarity.

Most of this work has been conducted on major depressive disorder (MDD). Some studies reported that participants ascertained using single-item measures of diagnosis have high genetic overlap with algorithm-based or clinically-ascertained MDD samples (Howard et al., 2019; Wray et al., 2018), suggesting comparability between individuals ascertained with the two measures. However, algorithm-based MDD has been found to have significantly higher heritability than single-item MDD. Higher heritability means more power to detect significant genetic effects. The higher heritability of algorithm-based MDD suggests that there are differences between algorithm-based and single-item diagnoses, and also implies that utilising the single-item measure could decrease the power to detect genetic effects (Cai et al., 2020; Glanville et al., 2020). If the two methods are not comparable, then measure selection or meta-analyses across cohorts with different ascertainment methods may impact the detection of genetic, as well as other (e.g., demographic, environmental, social), risk factors and outcomes. However, if instead the two methods agree well, not only does this support meta-analyses across datasets using these two approaches, it also reduces the burden on future participants, researchers, and clinicians in ascertaining diagnoses. Understanding the agreement between these measures is an important goal with clear implications across research and clinical fields.

In this study, we compared algorithm-based and single-item lifetime diagnoses for MDD and the five core anxiety disorders (generalised anxiety disorder [GAD], specific phobia, social anxiety disorder, panic disorder, and agoraphobia). Our aim was to assess agreement between these two diagnostic methods to determine whether they can be used interchangeably in research.

## Methods

### Sample

Data were examined from the National Institute for Health Research (NIHR) BioResource cohort (N = 59,161).

This included 41,708 participants that had been recruited as part of the Genetic Links to Anxiety and Depression (GLAD) Study (https://gladstudy.org.uk). The GLAD Study is an online research platform to recruit individuals with a lifetime experience of anxiety and/or depression for future research. Recruitment began in September 2018 and was conducted via traditional and social media campaigns or participating NHS sites.

The remaining 17,453 participants were NIHR BioResource members that had taken part in the COVID-19 Psychiatry and Neurological Genetics study (COPING NBR). This included members of the Irritable Bowel Disease cohort (IBD; N = 3,313) and general population cohorts (N = 14,140). Initial recruitment to the NIHR BioResource occurred in a variety of ways, including through blood donation centres.

Both studies were conducted entirely online. Eligibility was limited to those aged 16 and over and who lived in the UK. Eligibility for the GLAD Study also required a lifetime experience of an anxiety or depressive disorder. Assessment occurred at a single stage in which all participants responded to online, self-report questionnaires that included two methods for ascertaining likely depressive and anxiety disorder diagnoses: algorithm-based and single-item. The analyses presented in this paper include data from all participants that completed the GLAD or COPING survey before December 10th, 2020. Additional details of the design and implementation of the GLAD Study are described elsewhere (14).

### Measures

#### Algorithm-based diagnoses

*Algorithm*-*based* diagnoses were evaluated using the MDD, GAD, specific phobia, social anxiety disorder, panic disorder, and agoraphobia modules from an adapted version of the short form Composite International Diagnostic Interview (CIDI-SF) (15), as used in the UK Biobank (16) and Australian Genetics of Depression study (17). The CIDI-SF is based on the DSM-5 criteria for the disorders. Some validation studies of the self-report CIDI-SF for MDD have shown comparable agreement between algorithm-based MDD with diagnostic interviews (18,19). However, another study found low agreement between the self-report CIDI-SF for all disorders (MDD, GAD, specific phobia, social anxiety disorder, panic disorder, and agoraphobia) and structured interviews (20). Algorithms were developed to categorise participants as having a lifetime algorithm-based diagnosis for a disorder if their responses on the CIDI-SF corresponded closely to DSM-5 criteria (see Appendix 1 in Supplementary Materials).

#### Single-item diagnoses

*Single-item* diagnoses were self-reported responses to the question: “Have you ever been diagnosed with one or more of the following mental health problems by a professional, even if you don’t have it currently?” Participants were prompted to select all diagnoses that applied or indicate “None of the above”. Participants that did not respond to the single-item measure therefore had missing data for all single-item diagnoses. For the GLAD cohort, single-item panic disorder was added partway through data collection and is only included for GLAD participants that responded after 5 November 2018. Participants that signed up before that date were excluded from all agreement analyses for panic disorder. We included single-item “panic attacks” as well as “panic disorder”, and separately compared both to algorithm-based panic disorder. We are mindful that panic attacks are transdiagnostic and not specific to panic disorder. Research has shown that patients who have a panic attack are more likely to seek help from physical health professionals (e.g. in hospitals) rather than mental health services (21,22). However, recognition and diagnosis of panic disorder from physical health professionals is low (23,24). Given the higher recognition of panic attacks compared to panic disorder, we were interested in comparing agreement between single-item diagnosis of panic attacks and panic disorder with algorithm-based panic disorder. Participants were categorised as having a single-item diagnosis if they selected the most comparable option to the relevant diagnosis (e.g., “Depression” for MDD). Phrasing for each of these items can be found in Appendix 2 in Supplementary Materials. These single-item diagnoses reflect self-reports of a previous medically-provided diagnosis and were not validated against electronic health records (EHR). Validation studies for single-item diagnoses have found moderate agreement of single-item MDD (25,26) but poor agreement for single-item anxiety disorders (27) with structured interviews.

#### “Any anxiety” diagnosis

It is common in research to combine the anxiety disorder subtypes into a single category, given that the risk factors and outcomes overlap considerably between them (e.g., Purves et al., 2019 (28)). We were interested in assessing agreement of algorithm-based and single-item diagnoses of “any anxiety” as well as that for the individual anxiety disorders. Algorithm-based “any anxiety disorder” was defined as participants with an algorithm-based diagnosis for at least one of the individual anxiety disorders (e.g., GAD, specific phobia, social anxiety disorder, panic disorder, or agoraphobia). Single-item diagnosis of “any anxiety disorder” included participants who self-reported receiving at least one anxiety disorder diagnosis from a health professional.

### Analysis

We calculated the number of participants with zero, one, and two or more algorithm-based and single-item diagnoses. Participants with at least one missing value for an algorithm-based diagnosis were included in the frequencies for one, two, or three or more algorithm-based diagnoses. However, they were excluded when calculating the number of participants with zero algorithm-based diagnoses. Single-item panic disorder was added partway through GLAD data collection resulting in 14,858 GLAD participants with missing data on this item. These participants with missing data on single-item panic disorder were included in all single-item disorder frequencies. Participants with missing data on the remaining single-item diagnoses were excluded.

We also assessed the frequency of algorithm-based and single-item diagnoses for each disorder as percentages of the whole sample, excluding participants with missing data on one of the measures for the disorder in question (e.g., a participant with single-item GAD but missing data for algorithm-based GAD was excluded from the GAD frequencies).

Linear regression models were built to assess associations between demographic variables and missing data for algorithm-based, and logistic regressions were used to assess associations with single-item diagnoses.

Agreement and disagreement levels between these two diagnostic methods were assessed by calculating accuracy (the proportion [%] agreement), Cohen’s kappa (29), McNemar’s within-subjects chi-squared test (30), sensitivity, and specificity. Cohen’s kappa calculated reliability between the two methods. Values range from zero to one, with higher values indicating greater reliability (31). McNemar’s test assessed whether differences in the predictive accuracy of single-item and algorithm-based diagnoses were statistically significant (α < 0.05). Sensitivity is the proportion of individuals with a disorder that the measure correctly classifies as having a diagnosis (proportion of true positives). In contrast, specificity is the proportion of individuals without a disorder that are correctly classified as not having a diagnosis (proportion of true negatives). Since we lacked a ‘gold standard’ reference in this sample, sensitivity and specificity analyses were conducted in both directions. We interpreted results as “agreed positives/negatives” and “disagreed positives/negatives”.

The proportions of agreement and disagreement between these measures were also examined post-hoc by sex and compared using chi-squared analyses.

### Code availability

All data cleaning and analyses were conducted using R version 3.5.3 (32), the tidyverse (33), and caret (34) packages. The full code for the diagnostic algorithms and analyses included in this paper are available at https://github.com/mollyrdavies/GLAD-Diagnostic-algorithms.

### Data availability

The data that support the findings of this study are available on request from the corresponding author, TCE. The data are not publicly available due to restrictions outlined in the study protocol and specified to participants during the consent process.

## Results

### Sample characteristics

Participants with missing data for sex (N = 754 GLAD only) or age (N = 31 GLAD; N = 6 COPING NBR) were excluded from analyses. The remaining sample included 58,400 participants. Table 1 displays the sample descriptives by cohort. The average age of participants was 43 years, 73% were female, the majority self-defined as white (95%), and a large proportion had a university degree (54%). Characteristics between the cohorts were compared with t-test and chi-squared analyses. Given the large sample sizes, all characteristics were significantly different between the cohorts. The only differences that were clinically meaningful were age and sex, with the GLAD sample having a younger mean age and a higher proportion of female participants.

**Table 1.**
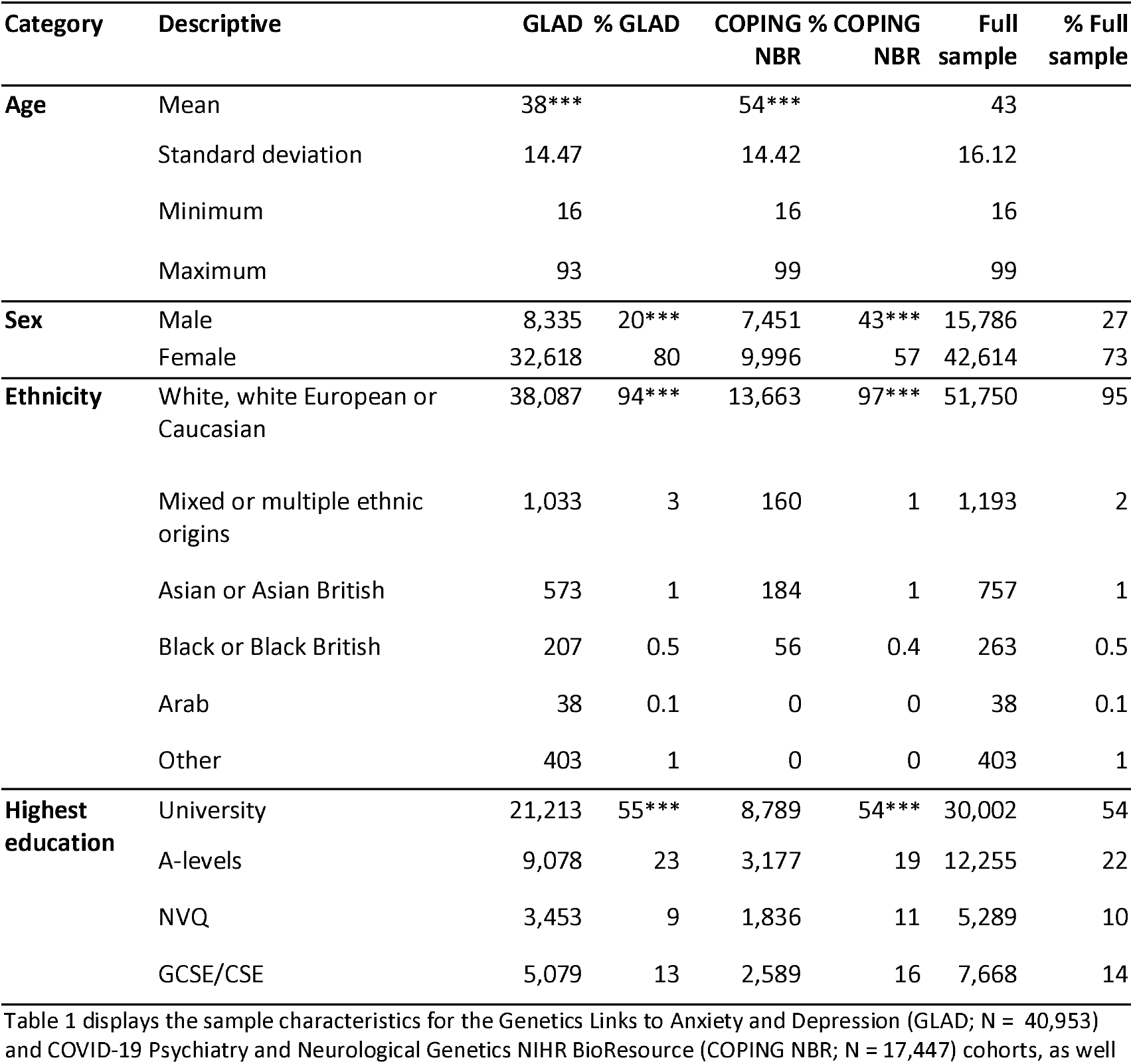
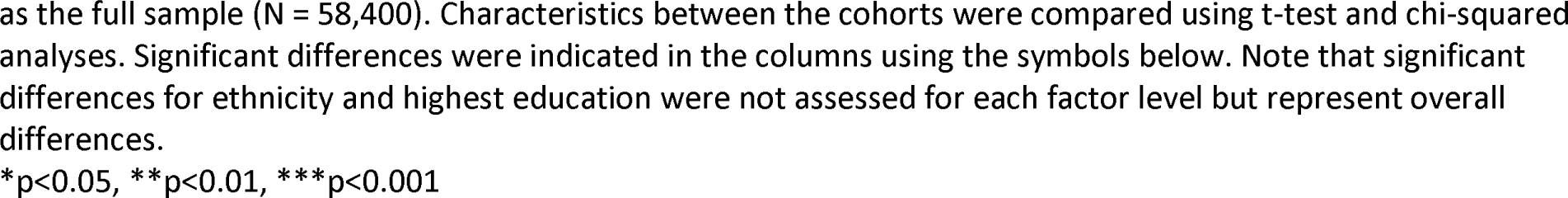
Sample characteristics.

| Category | Descriptive | GLAD | % GLAD | COPING NBR | % COPING NBR | Full sample | % Full sample |
| --- | --- | --- | --- | --- | --- | --- | --- |
| **Age** | Mean | 38*** |  | 54*** |  | 43 |  |
|  | Standard deviation | 14.47 |  | 14.42 |  | 16.12 |  |
|  | Minimum | 16 |  | 16 |  | 16 |  |
|  | Maximum | 93 |  | 99 |  | 99 |  |
| **Sex** | Male | 8,335 | 20*** | 7,451 | 43*** | 15,786 | 27 |
|  | Female | 32,618 | 80 | 9,996 | 57 | 42,614 | 73 |
| **Ethnicity** | White, white European or Caucasian | 38,087 | 94*** | 13,663 | 97*** | 51,750 | 95 |
|  | Mixed or multiple ethnic origins | 1,033 | 3 | 160 | 1 | 1,193 | 2 |
|  | Asian or Asian British | 573 | 1 | 184 | 1 | 757 | 1 |
|  | Black or Black British | 207 | 0.5 | 56 | 0.4 | 263 | 0.5 |
|  | Arab | 38 | 0.1 | 0 | 0 | 38 | 0.1 |
|  | Other | 403 | 1 | 0 | 0 | 403 | 1 |
| **Highest education** | University | 21,213 | 55*** | 8,789 | 54*** | 30,002 | 54 |
|  | A-levels | 9,078 | 23 | 3,177 | 19 | 12,255 | 22 |
|  | NVQ | 3,453 | 9 | 1,836 | 11 | 5,289 | 10 |
|  | GCSE/CSE | 5,079 | 13 | 2,589 | 16 | 7,668 | 14 |

Table 1 displays the sample characteristics for the Genetics Links to Anxiety and Depression (GLAD; N = 40,953) and COVID-19 Psychiatry and Neurological Genetics NIHR BioResource (COPING NBR; N = 17,447) cohorts, as well
as the full sample (N = 58,400). Characteristics between the cohorts were compared using t-test and chi-squared analyses. Significant differences were indicated in the columns using the symbols below. Note that significant differences for ethnicity and highest education were not assessed for each factor level but represent overall differences.
\*p<0.05, \*\*p<0.01, \*\*\*p<0.001

### Frequencies

The frequency of the number of major depressive or anxiety disorder diagnoses was examined (Table 2). Since panic attacks are not a disorder, they were excluded from single-item disorder frequencies. For algorithm-based diagnoses, 21,779 participants (13,486 GLAD and 8,313 COPING NBR) had missing data for at least one disorder. Participants with zero single-item or algorithm-based diagnoses that have missing data for at least one disorder on the respective measure (excluding single-item panic disorder, which was added partway through data collection) are displayed in Table 2 in the appropriate “NA” column.

**Table 2.** Frequencies of algorithm-based and single-item diagnoses from the full sample (N = 58,400) and by cohort.

|  |  | Algorithm-based diagnoses |  |  |  |  |  | Single-item diagnoses<sup>b</sup> |  |  |  |  |  |
| --- | --- | --- | --- | --- | --- | --- | --- | --- | --- | --- | --- | --- | --- |
| Subset | Total | 0 | 1 | 2 | 3+ | NA | Mean | 0 | 1 | 2 | 3+ | NA | Mean |
| Full sample | 58,400 | 7,395<br>(13%) | 13,461<br>(23%) | 10,570<br>(18%) | 17,721<br>(30%) | 9,253<br>(16%) | 1.78 | 14,268<br>(24%) | 11,755<br>(20 %) | 20,032<br>(34%) | 10,700<br>(18%) | 1,645<br>(3%) | 1.54 |
| GLAD | 40,953 | 1,158<br>(3%) | 9,867<br>(24%) | 9,296<br>(23%) | 17,234<br>(42%) | 3,398<br>(8%) | 2.34 | 1,631<br>(4%) | 8,725<br>(21.3%) | 18,736<br>(46%) | 10,357<br>(25%) | 1,504<br>(4%) | 2.04 |
| COPING NBR | 17,447 | 6,237<br>(36%) | 3,594<br>(21%) | 1,274<br>(7%) | 487<br>(3%) | 5,855<br>(34%) | 0.45 | 12,637<br>(72%) | 3,030<br>(17%) | 1,296<br>(7%) | 343<br>(2%) | 141 (1%) | 0.39 |
| One or more<br>algorithm-based diagnosis | 41,752 |  |  |  |  |  |  | 3,322<br>(8%) | 9,435<br>(21%) | 18,567<br>(44%) | 10,223<br>(24%) | 205<br>(1%) | 1.94 |
| One or more<br>single-item<br>diagnosis | 42,487 | 1,460<br>(3%) | 11,138<br>(26%) | 9,830<br>(23%) | 17,257<br>(41%) | 2,802<br>(7%) | 2.32 |  |  |  |  |  |  |

The table displays the number and percentage of the full sample, GLAD, and COPING NBR participants (rows) with 0, 1, 2, or 3+ algorithm-based or single-item diagnoses (columns). The mean number of algorithm-based and single-item diagnoses are reported. We also report the frequencies with 0, 1, 2, or 3+ algorithm-based or single-item diagnoses for participants with one or more diagnoses for the opposing measure (e.g., the number of participants with one or more algorithm-based diagnoses who have 0, 1, 2, or 3+ single-item diagnoses).
<sup>a</sup>For algorithm-based diagnoses, 21,779 participants (13,486 GLAD and 8,313 COPING NBR) had missing data for at least 1 disorder. Participants with at least 1 missing value for an algorithm-based diagnosis were excluded when calculating the number of participants with 0 algorithm-based diagnoses, and are instead included in the “NA” column under the “algorithm-based diagnoses” header. Participants with missing data were included in the remaining frequencies for 1, 2, or 3+ algorithm-based diagnoses.
<sup>b</sup>For single-item diagnoses, the panic disorder was added partway through data collection. Participants with missing data on any single-item diagnosis except panic disorder were excluded from these frequencies, but those with missing data for panic disorder were included. The number of excluded participants are presented in the “NA” column under the “single-item diagnoses” header. Panic attacks were not included in these figures.
Abbreviations: GLAD, Genetics Links to Anxiety and Depression; COPING NBR, COVID-19 Psychiatry and Neurological Genetics NIHR BioResource

Being male, self-identifying as Mixed or Asian/Asian British, and having a lower level of educational attainment were significantly associated with more missing data on algorithm-based diagnoses. No characteristics were meaningfully associated with missing data on single-item diagnoses. Full details of these analyses and summary of the results can be found in Appendix 3 in Supplementary Materials, along with a table of missing data by diagnosis and frequencies of the number of missing algorithm-based diagnoses.

Frequency of diagnosis varied by cohort. As shown in Table 2, of GLAD participants only 3% did not report an algorithm-based diagnosis and 4% did not report a single-item diagnosis for any disorder. However, for the COPING NBR sample these proportions were greater with 36% without an algorithm-based and 72% without a single-item diagnosis. Of note, the proportion of COPING NBR participants without an algorithm-based diagnosis was lower due to the high percentage (34%) in the missing data (NA) column, which included participants with no algorithm-based diagnosis and missing data on the algorithm for at least one disorder. The difference in proportions between the cohorts is unsurprising, since GLAD participants identified themselves as having had an anxiety and/or depressive diagnosis at some point in their lives whereas COPING NBR participants were recruited from the general population or for a physical health condition. Overall, 42,487 (73%) participants reported a single-item diagnosis of a major depressive or anxiety disorder, whereas 41,752 (71%) participants were identified as having at least one of the algorithm-based diagnoses.

Figure 1 displays the frequencies of algorithm-based and single-item diagnoses for the full sample and by cohort for each of the disorders. The bars for each diagnosis only include participants without missing data for either measure on the specified disorder. For instance, the proportion of participants with a MDD diagnosis was calculated as a percentage of those with no missing data for neither algorithm-based nor single-item MDD.

**Figure 1.**
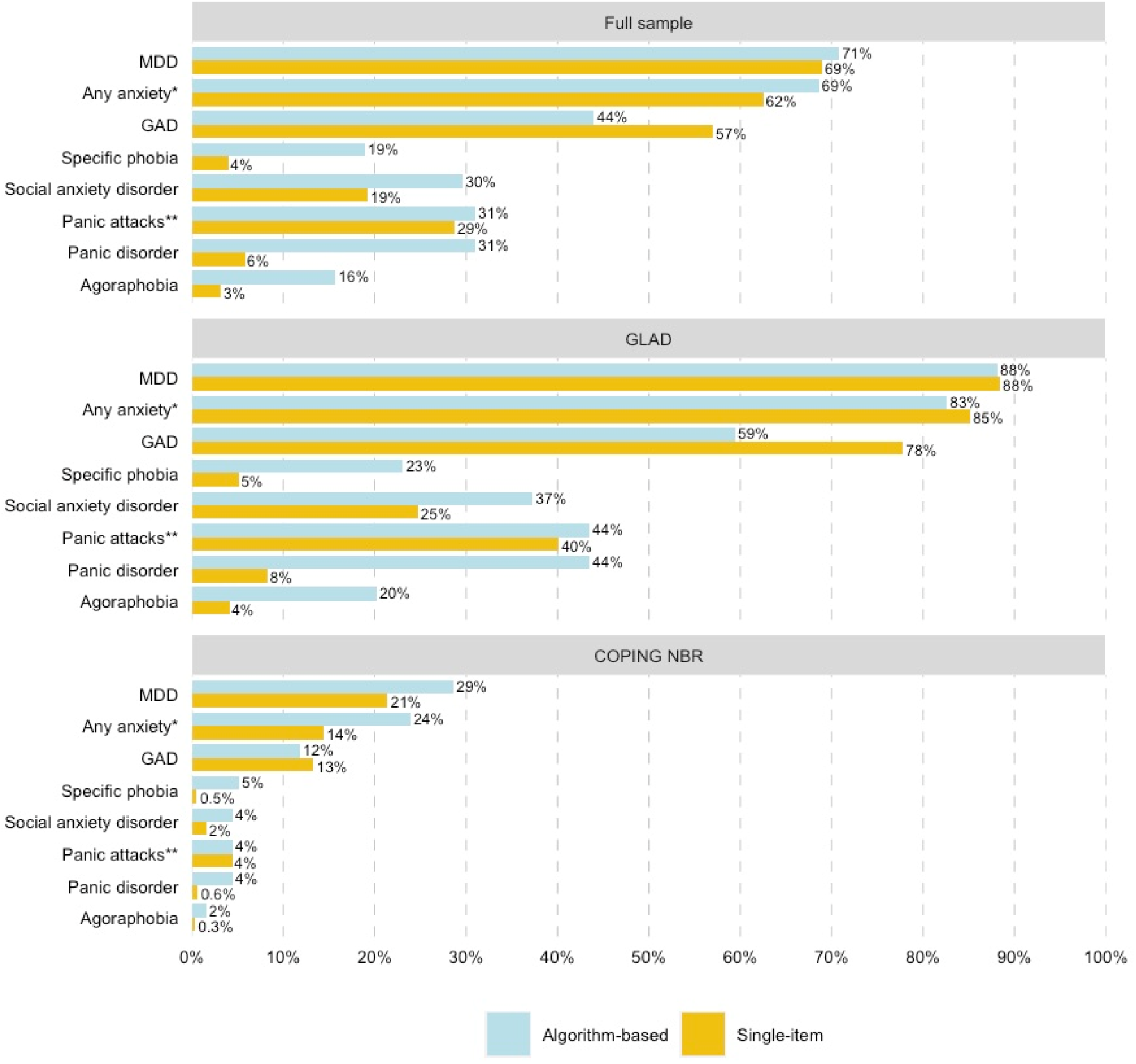
Frequencies of algorithm-based and single-item diagnoses of major depressive disorder, any anxiety, or an anxiety disorder in the GLAD and COPING cohorts. The bars represent the proportion (%) of either the full sample (N = 58,400), GLAD (N = 40,953), or COPING NBR (N = 17,447) with an algorithm-based (blue) or single-item diagnosis (yellow) for each disorder. Proportions exclude participants with missing data on either measure for the specified diagnosis. ^*^Any anxiety includes participants with at least one anxiety disorder (GAD, specific phobia, social anxiety disorder, panic disorder, and/or agoraphobia) on the indicated method (algorithm-based vs single-item). ^* *^For panic attacks, algorithm-based panic disorder is displayed and compared to single-item panic attacks. Abbreviations: GLAD, Genetics Links to Anxiety and Depression; COPING NBR, COVID-19 Psychiatry and Neurological Genetics NIHR BioResource; MDD, major depressive disorder; GAD, generalised anxiety disorder

MDD had the highest frequency for both cohorts, which was relatively consistent across diagnostic methods (GLAD: 88% algorithm-based, 88% single-item; COPING NBR: 29% algorithm-based, 21% single-item). The frequencies of the anxiety disorders varied widely depending on cohort and measure. The majority of GLAD participants had a single-item diagnosis of GAD (78%) but the percentage with an algorithm-based GAD diagnosis (59%) was approximately two-thirds of that value. In contrast, the percentage of COPING NBR participants with algorithm-based (12%) and single-item (13%) GAD was similar. The remaining anxiety disorders, which consisted of the phobic disorders, had higher frequencies of algorithm-based than single-item diagnoses. For instance, the percentages of participants with algorithm-based specific phobia (GLAD: 23%; COPING NBR: 5%), panic disorder (GLAD: 44%; COPING NBR: 5%), and agoraphobia (GLAD: 20%; COPING NBR: 2%) were more than double those of the respective single-item diagnoses (GLAD: 4-8%; COPING NBR: 0.3-0.6%). The proportion of participants with algorithm-based panic disorder (GLAD: 44%; COPING NBR: 5%) was more similar to single-item panic *attacks* (GLAD: 40%; COPING NBR: 4%) than single-item panic disorder (GLAD: 8%; COPING NBR: 0.6%).

### Agreement

We examined the agreement between algorithm-based and single-item diagnoses. Figure 2 displays the agreement and disagreement for each disorder. Accuracy, sensitivity, specificity, Cohen’s kappa, and McNemar’s test p-values are presented in Table 3.

**Figure 2.**
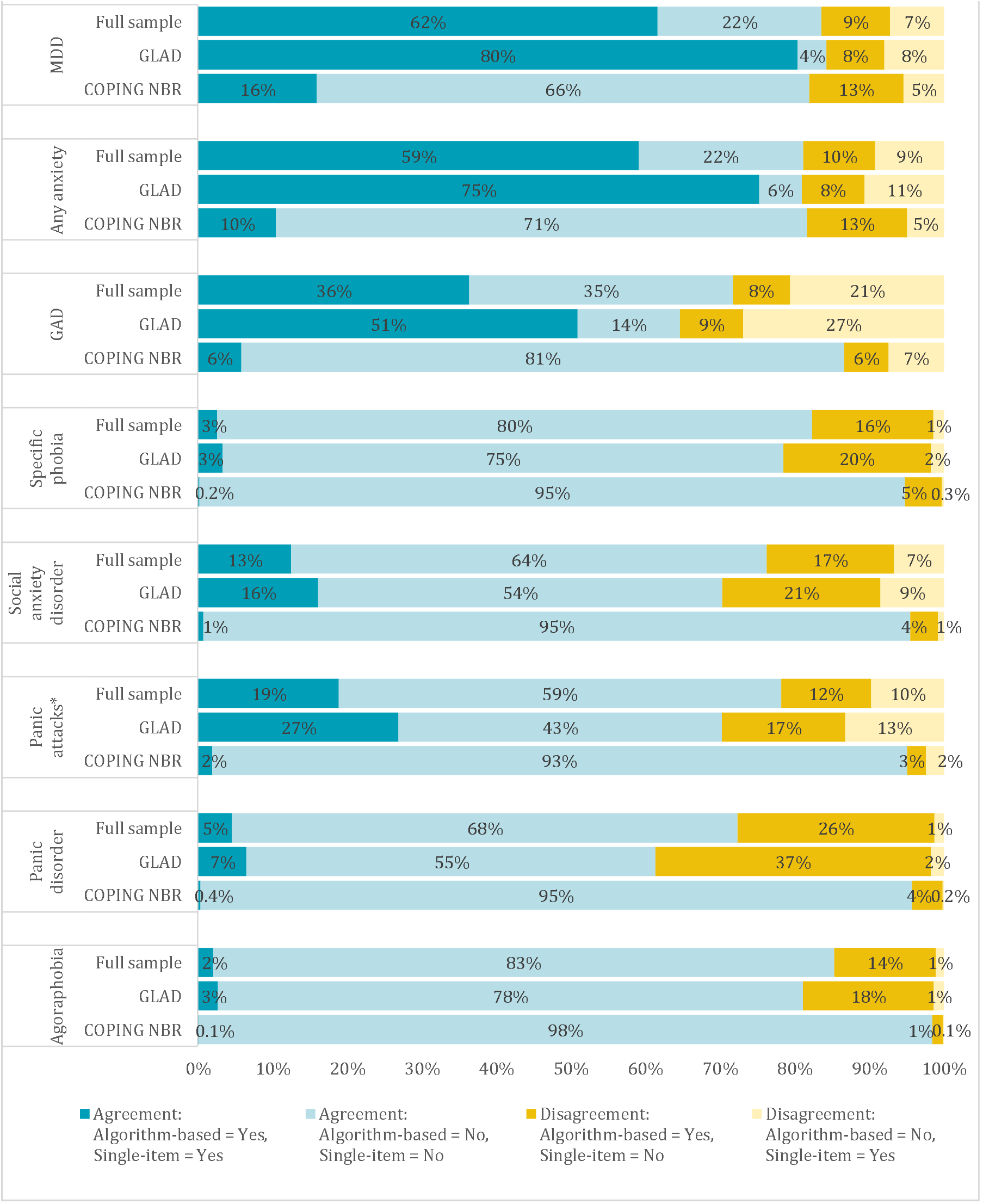
All comparisons of agreement and disagreement on algorithm-based vs single-item diagnoses. Each bar displays the proportions (%) of the full sample (N = 58,400) and each cohort (GLAD: N = 40,954; COPING NBR: N = 17,447) with agreement or disagreement between the two measures for each disorder. Agreements are represented in blue (dark blue = agreement on diagnosis, light blue = agreement on no diagnosis) while disagreements are in yellow (dark yellow = algorithm-based but not single-item diagnosis, light yellow = single-item but not algorithm-based diagnosis). ^*^The panic attacks column displays the agreement between algorithm-based panic disorder and single-item panic attacks. Abbreviations: GLAD, Genetics Links to Anxiety and Depression; COPING NBR, COVID-19 Psychiatry and Neurological Genetics NIHR BioResource; MDD, major depressive disorder; GAD, generalised anxiety disorder

**Table 3.**
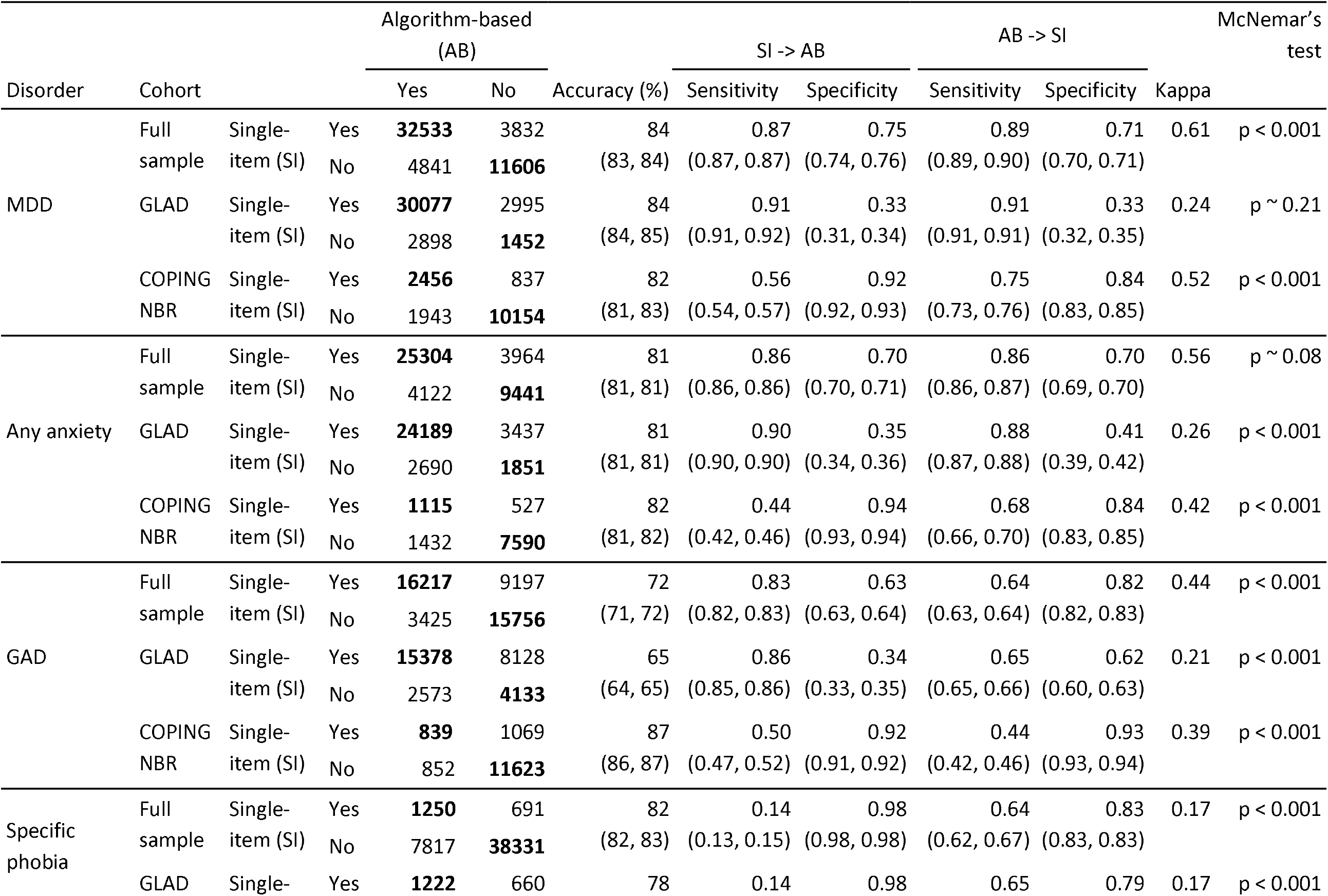
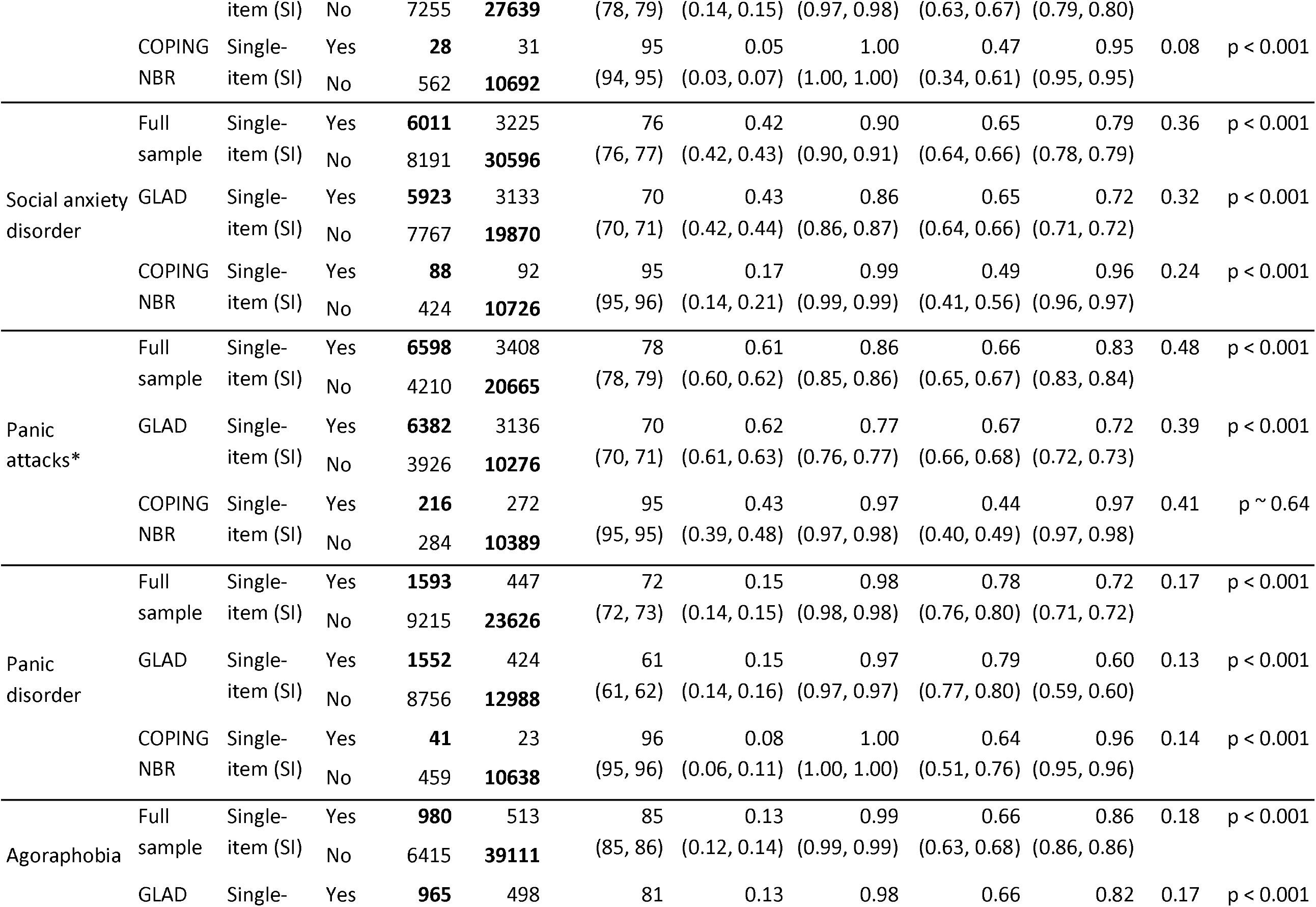
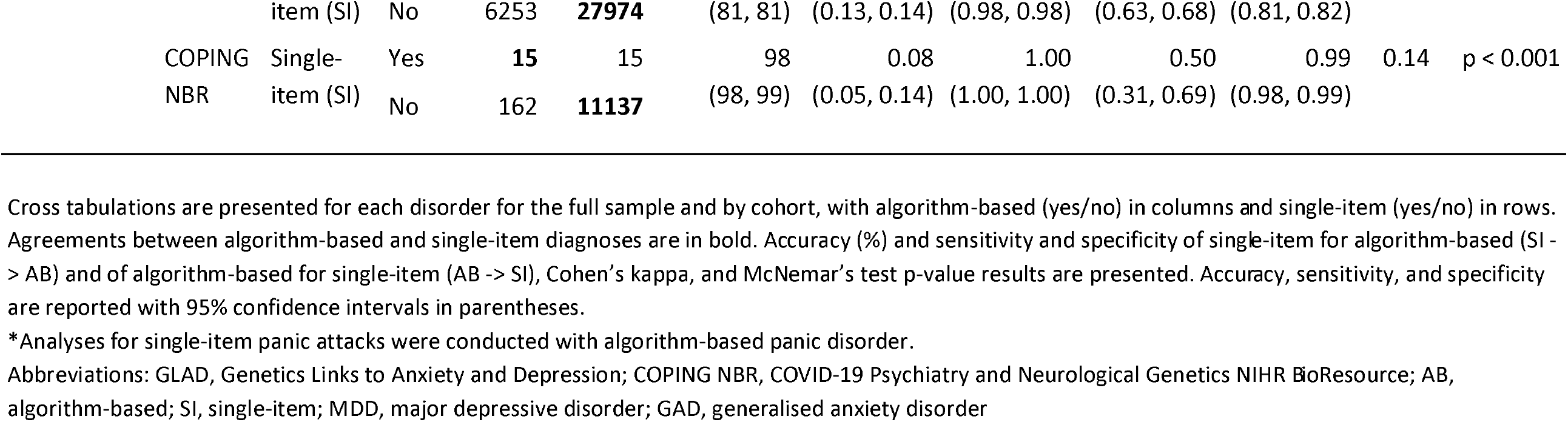
Agreement between algorithm-based and single-item diagnoses for the full sample and by cohort.

We see a fairly similar pattern of effects for MDD, any anxiety, and GAD in the full sample. The accuracy or proportion (%) agreement between algorithm-based and single-item diagnoses for these disorders was high (72-84%). Similarly, single-item diagnoses had high sensitivity (0.83-0.87) and moderate specificity (0.63-0.75) for the respective algorithm-based measure. This indicates that these *single-item* diagnoses of MDD, any anxiety, and GAD had high proportions of agreed positives and slightly lower proportions of agreed negatives with the algorithm-based measure. Notably, sensitivity and specificity of *algorithm-based* MDD and any anxiety for single-item diagnoses was similar, meaning that proportions of agreed positives and agreed negatives for these disorders were comparable between these measures, regardless of the direction of comparison. In contrast, algorithm-based GAD had moderate sensitivity and high specificity for the single-item measure. Single-item GAD in the full sample therefore had higher sensitivity (e.g., proportion of agreed positives) for the algorithm-based diagnosis than algorithm-based GAD had for the single-item diagnosis. These sensitivity results correspond with Figure 2, which demonstrated that the largest proportion of disagreement for GAD (21%) were participants with a single-item but not algorithm-based diagnosis. Instead, MDD and any anxiety had equal proportions of the two types of disagreement, hence sensitivity and specificity results for these measures were similar in both directions. Yet despite the reasonable sensitivity and specificity values for MDD, any anxiety, and GAD in the full sample, Cohen’s kappa indicated only moderate agreement for these measures (31).

When results for these three disorder categories are broken down by cohort, the proportion of agreement for MDD and any anxiety remained high for both GLAD and COPING NBR (81-84%), while the proportion of agreement for GAD was lower in GLAD (65%) than COPING NBR (87%). However, sensitivity and specificity results varied by cohorts. In the GLAD cohort, single-item MDD, any anxiety, and GAD had high sensitivity (0.86-0.91) and low specificity (0.33-0.35) for the algorithm-based measures, whereas these single-item diagnoses in the COPING NBR cohort had low to moderate sensitivity (0.44-0.56) and high specificity (0.92-0.94). For example, single-item MDD had sensitivity of 0.91 and specificity of 0.33 in GLAD but sensitivity of 0.56 and specificity of 0.92 in COPING NBR. Single-item MDD, any anxiety, and GAD therefore had high proportions of agreed positives in GLAD, and high proportions of agreed negatives in COPING NBR. Differences were observed in sensitivity and specificity results of the algorithm-based diagnoses for the single-item measures as well, with GLAD having higher sensitivity and lower specificity on each disorder than COPING NBR. For instance, algorithm-based MDD had sensitivity of 0.91 and specificity of 0.33 in GLAD and sensitivity of 0.75 and specificity of 0.84 in COPING NBR for single-item MDD.

Single-item diagnoses of the phobic disorders (specific phobia, social anxiety disorder, panic disorder, and agoraphobia) showed a consistent pattern of results in the full sample and by cohort, displaying low sensitivity (0.08-0.43) and high specificity (0.86-1.00) for the algorithm-based measures. Referring to Figure 2, the highest proportion of disagreement between the phobic disorder diagnoses was observed for participants with an algorithm-based but not single-item diagnosis. These results were more pronounced for the COPING NBR cohort, for which the single-item diagnoses of the phobic disorders had sensitivity values below 0.17 and specificity around 1.00 for the algorithm-based measure. Although the proportion of agreement for these disorders ranged between 61% (panic disorder in GLAD) and 98% (agoraphobia in COPING NBR), Cohen’s kappa values indicated slight agreement for specific phobia, panic disorder, and agoraphobia diagnoses and fair agreement for social anxiety disorder in the full sample and by cohort.

The single-item measure of panic attacks had higher sensitivity and lower specificity than single-item panic disorder for algorithm-based panic disorder in the full sample and by cohort. For example, in the full sample single-item panic attacks had sensitivity of 0.61 and specificity of 0.86, while single-item panic disorder had sensitivity of 0.15 and specificity of 0.98 for algorithm-based panic disorder. Single-item panic attacks therefore had a higher proportion of agreed positives but lower proportion of agreed negatives than single-item panic disorder for the algorithm-based measure, also observable in Figure 2. Algorithm-based panic disorder showed a comparable proportion of agreement with single-item panic attacks and panic disorder, but Cohen’s kappa indicated better agreement with single-item panic attacks.

### Sex differences in agreement

There was little variation by sex observed in proportions of agreement and disagreement between the measures (Appendix 4 in Supplementary Materials).

### Alternate diagnoses

There was an unusually large proportion of the full sample, relative to the other disorders, who reported a single-item diagnosis of GAD but did not receive an algorithm-based diagnosis (21%). We conducted a post-hoc analysis to explore whether these participants received any other algorithm-based diagnosis. Figure 3 displays the proportions (%) of participants with single-item but not algorithm-based GAD that had other algorithm-based diagnoses, as well as the proportion without any algorithm-based diagnosis. Proportions were calculated excluding participants with missing data for that disorder. The proportion of participants with no algorithm-based diagnosis excluded participants with missing data on any of the disorders.

**Figure 3.**
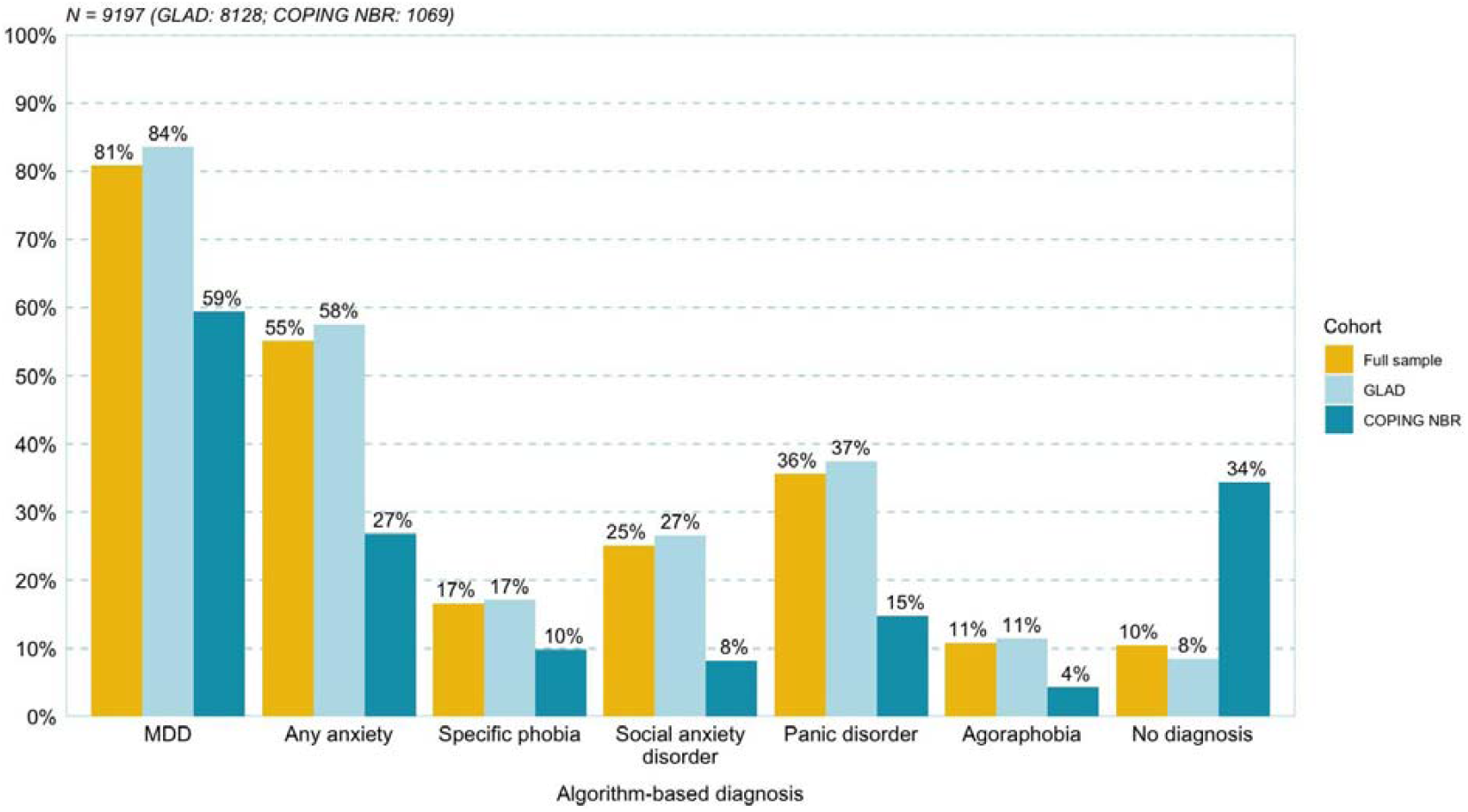
Alternate algorithm-based diagnoses for participants with single-item but not algorithm-based generalised anxiety disorder. Each bar displays the proportions (%) of participants with single-item but not algorithm-based GAD from the full sample (N = 9,197) and each cohort (GLAD: N = 8,128; COPING NBR: N = 1,069) with algorithm-based diagnoses for each of the other disorders, for any other algorithm-based anxiety disorder, or without any algorithm-based diagnosis (no diagnosis). The bars for the disorders are not exclusive as participants may have more than one algorithm-based diagnosis. Proportions exclude participants with missing data for the respective single-item diagnosis, and the proportion for “no diagnosis” excludes participants with missing data on any of the disorders. Abbreviations: GLAD, Genetics Links to Anxiety and Depression; COPING NBR, COVID-19 Psychiatry and Neurological Genetics NIHR BioResource; MDD, major depressive disorder; GAD, generalised anxiety disorder

The results showed that the largest proportion of participants with single-item but not algorithm-based GAD for both cohorts had algorithm-based MDD (GLAD: 84%; COPING NBR: 59%). Of the anxiety disorders, algorithm-based panic disorder was the most common for these participants (GLAD: 37%; COPING NBR: 15%). Over half of GLAD participants with single-item but not algorithm-based GAD had a different algorithm-based anxiety disorder (58%), with only 8% having no algorithm-based diagnosis. For COPING NBR, approximately a quarter of these participants had a different algorithm-based anxiety disorder (27%), but over one-third did not have any algorithm-based diagnosis (34%).

The phobic disorders (specific phobia, social anxiety disorder, panic disorder, and agoraphobia) displayed relatively high proportions of participants with algorithm-based but not single-item diagnoses. For these disorders, we were interested in exploring whether these participants reported other single-item diagnoses, different from the algorithm-based diagnosis they received. We hypothesised that a large number of these participants would report a single-item GAD diagnosis. Figure 4 displays the proportions of participants with single-item but not algorithm-based diagnoses for specific phobia (N = 7,817), social anxiety disorder (N = 8,191), panic disorder (N = 9,215), and agoraphobia (N = 6,415) who reported a single-item diagnosis for another disorder.

**Figure 4.**
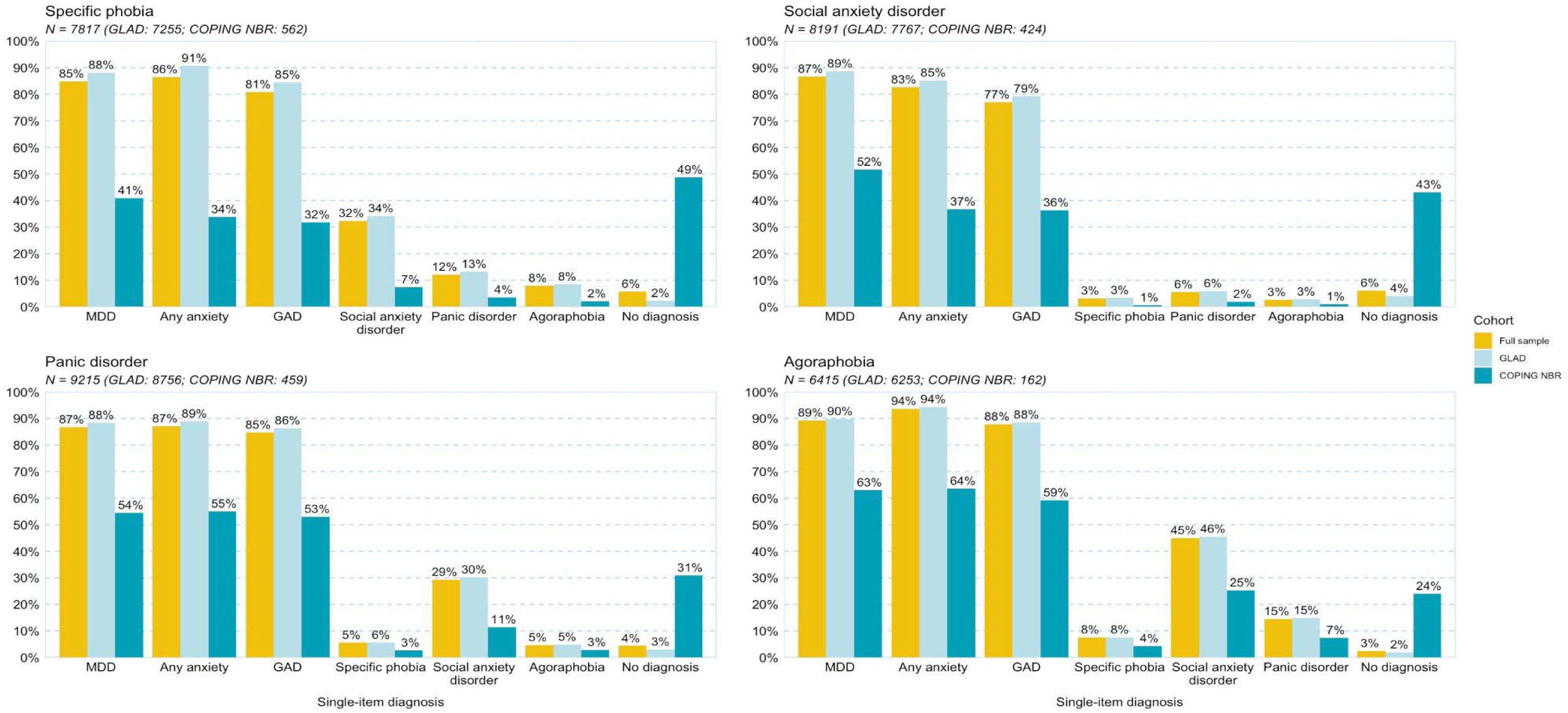
Alternate single-item diagnoses for participants with algorithm-based but not single-item specific phobia, social anxiety disorder, panic disorder, or agoraphobia. Each graph includes participants with an algorithm-based but not single-item diagnosis of specific phobia, social anxiety disorder, panic disorder, or agoraphobia. The titles of the plots indicate the disorder, and subtitles display the N of participants from the full sample and each cohort with a single-item and no algorithm-based diagnosis for the referenced disorder. The bars display the proportions (%) of these participants with single-item diagnoses for each of the other disorders, for any other single-item anxiety disorder, or without any single-item diagnosis (no diagnosis). The bars for the disorders are not exclusive as participants may have more than one single-item diagnosis. Proportions exclude participants with missing data for the respective single-item diagnosis, and the proportion for “no diagnosis” excludes participants with missing data on any of the disorders. Abbreviations: GLAD, Genetics Links to Anxiety and Depression; COPING NBR, COVID-19 Psychiatry and Neurological Genetics NIHR BioResource; MDD, major depressive disorder; GAD, generalised anxiety disorder

Those with an algorithm-based but not single-item diagnosis of the phobic disorders all showed a somewhat similar pattern of alternative diagnoses. Specifically, MDD and GAD were the most common alternate single-item diagnoses, reported by over three-quarters of the GLAD cohort and between one-third and half of the COPING NBR cohort. Single-item social anxiety disorder was also reported by over one-third of the GLAD cohort. For COPING NBR, in those with algorithm-based but not single-item agoraphobia, only social anxiety disorder was reported at a relatively increased rate. Alternate single-item diagnoses of the phobic disorders were rarely reported in either cohort. Notably, between one-quarter and half of the COPING NBR cohort with an algorithm-based but single-item diagnosis of one of these phobic disorders did not report a single-item diagnosis of any disorder, which was highly uncommon in the GLAD cohort (less than 4%).

## Discussion

### Overview

In this study, we examined the agreement and disagreement between algorithm-based and single-item lifetime diagnoses of MDD, any anxiety, and the five core anxiety disorders: GAD, specific phobia, social anxiety disorder, panic disorder, and agoraphobia. These approaches are often utilised as the sole diagnostic method in large-scale research studies and frequently combined in meta-analyses despite limited evidence regarding comparability.

Algorithm-based and single-item diagnoses for MDD and any anxiety were reasonably comparable, demonstrating high accuracy (81-84%) and moderate agreement (κ = 0.56-0.61). Particularly high agreement was found for participants *with* a diagnosis (sensitivity 0.86-0.87). Although accuracy (72%) and Cohen’s kappa (κ = 0.44) indicated moderate agreement for the GAD measures, a large proportion (21%) of the sample with single-item GAD did not receive an algorithm-based diagnosis, an effect not seen for MDD or any anxiety. This parallels findings from 150,000 individuals recruited from the general population into the UK Biobank, which also found moderate agreement for MDD (κ = 0.46) but much lower agreement for GAD (κ = 0.28). Interestingly, the single-item measures for MDD, any anxiety, and GAD performed well at identifying *algorithm-based cases* in the GLAD case cohort (sensitivity 0.86-0.90), but poorly in COPING NBR (sensitivity 0.44-0.56). In contrast, this approach performed well at identifying *algorithm-based controls* in the COPING NBR general population cohort (specificity 0.92-0.94) yet was poor at identifying those without a diagnosis in GLAD (specificity 0.33-0.35). This suggests that the enrichment of cases and controls in these two samples respectively had a large impact on the performance of the two approaches.

In contrast, for the phobic disorders (specific phobia, social anxiety disorder, panic disorder, and agoraphobia) we found only slight to fair agreement (κ = 0.17-0.36). Sensitivity and specificity results were also notably different for single-item assessment of the phobic disorders compared to MDD, any anxiety, and GAD. Single-item phobic disorder assessment displayed high agreement with the algorithm-based measure on participants *without* a diagnosis (specificity 0.90-0.99), but poor agreement on participants *with* a diagnosis (sensitivity 0.13-0.42). Post-hoc analyses found that a relatively large proportion of the participants (12-26%) in our sample that received an algorithm-based diagnosis of a phobic disorder did not report the same diagnosis, instead many reported single-item GAD or MDD.

Surprisingly, single-item panic attacks had higher sensitivity and only slightly lower specificity than single-item panic disorder for algorithm-based panic disorder. These findings are contrary to what might be expected, since panic attacks are a symptom that can manifest in isolation (35) and are not specific to panic disorder. However, single-item panic attacks captured a higher proportion of participants with algorithm-based panic disorder than single-item panic disorder.

### Limitations

A strength of the GLAD and COPING studies has been the successful recruitment of several thousand participants to complete detailed phenotyping measures. This enabled researchers to compare single-item and algorithm-based measures of depressive and anxiety disorders. However, as with any study, there are limitations that should be considered. Both cohorts are disproportionately female, white, and highly educated compared to the UK population. Exploration of measurement agreement in more representative samples may establish whether our findings are generalisable.

As mentioned previously, the algorithm-based and single-item diagnoses have not been compared to a ‘gold standard’ clinical interview in this study and prior evidence is conflicting (18–20,25–27). As a result, we cannot make any conclusions about which diagnostic method is more accurate from the analyses conducted here. Further research is therefore required to validate these measures against ‘gold standard’ clinical interviews. Validation is key to ensuring that research findings are relevant to clinical practice. Nonetheless, it is worth noting that some researchers have argued that a ‘gold standard’ diagnosis does not exist. Even structured and semi-structured interviews may result in different classifications of diagnosis and estimates of population prevalence (36). Other validation methods for these measures are worth exploring, such as investigating the genetic overlap with clinically-ascertained cohorts or by comparing against clinical outcome measures such as functional impairment or treatment response.

At this point we could not assess whether participants’ self-report of a clinical diagnosis matched their clinical data, nor which health professional provided the diagnosis (e.g., general practitioner [GP] or psychiatrist). In the context of genetics, studies which have utilised single-item diagnoses have similarly done so without medical record validation (28,37,38).

Furthermore, since individuals with depressive and anxiety disorders often do not present to a medical professional or receive a diagnosis (27,39,40), reliance on health records alone is not a substitute for asking the participant. However, all GLAD and COPING participants have consented to providing medical record access and an application for clinical data is underway, so this comparison could be conducted in future analyses.

### Implications

We observed an asymmetry in agreement results for MDD, any anxiety, and GAD, with agreement being better for cases in the case-enriched GLAD cohort and better for controls in the general population COPING NBR cohort. Consistent results across the cohorts were found for algorithm-based and single-item diagnoses of the phobic disorders (specific phobia, social anxiety disorder, panic disorder, and agoraphobia). The phobic disorder measures had high agreement for classification of participants *without* a diagnosis but differed substantially when classifying cases. Taken together, these findings suggest that studies on anxiety disorders applying single-item diagnostic methods would tend to categorise the majority of participants as having GAD, whereas those utilising algorithm-based measures would have a more even distribution across the anxiety subtypes.

These findings have important implications for large-scale studies investigating disorder-specific factors or outcomes, as ascertained diagnoses would differ depending on the measure. Although some factors are largely shared between major depressive and anxiety disorders (e.g., genetic factors), others show more specificity. For example, aspects of the environment show differential associations with depression and anxiety (41–43), and some suggest that incorporating disorder-specific approaches to psychological treatment can improve outcomes ((44)). Studies focussed on expanding sample sizes may find that meta-analyses combining data from cohorts ascertained with single-item or algorithm-based diagnoses is sufficient to identify effects that are shared between major depressive and anxiety disorders (42,45). However, in order to understand disorder-specific risk factors or investigate treatment approaches for these disorders, particularly the anxiety subtypes, findings may vary depending on the ascertainment method used in the study. The population of interest should therefore be considered when selecting measures for future studies. Both methods may have an important role in future research depending on the aims of individual studies. Those focussed on increasing participation and reducing the time burden for participants and researchers may consider the use of single-item measures, taking into account the differences by disorder in sensitivity and specificity for algorithm-based diagnoses. For instance, a study recruiting from clinical populations may use single-item MDD to ascertain MDD diagnosis. In contrast, those particularly interested in identifying cases with specific anxiety disorder subtypes are likely to benefit from use of the algorithm-based approach.

These results can further be considered in the context of the efficacy of single-item broad diagnostic measures, which have been explored the most thus far with regard to depression. There is a diversity of opinions as to the value and utility of these brief measures in the field, especially in large-scale research such as psychiatric genetics. For example, researchers have found that ‘broad’ depression (e.g., depression defined using single-item diagnoses or self-reports of treatment seeking) is non-specific to MDD and has lower heritability estimates than algorithm-based MDD (8,46). Misclassification dilutes the power of case-control analyses to detect differences between the samples (47,48). As such, the lower heritability estimates of single-item measures of MDD may indicate that this approach has a higher rate of misclassification for true cases and controls than the algorithm-based diagnoses. Indeed, single-item MDD, any anxiety, and GAD in this study had high agreement with algorithm-based diagnoses for identifying cases, but differed in the classification of those without a diagnosis. It has been argued that algorithm-based measures are preferable over the single-item counterpart, and that studies utilising single-item diagnoses for the purpose of case-control comparisons are more likely to identify effects that are non-specific, complicating efforts to disentangle disorder-specific factors and treatments (8,49). For studies that are unable to administer algorithm-based assessments or existing studies that did not include these measures, combining multiple broad diagnostic measures (e.g., single-item diagnoses, single-item help-seeking questions, and self-reported antidepressant usage) has been shown to reduce misclassification and increase heritability of MDD cases to equal or exceed heritability estimates of algorithm-based MDD (46).

In terms of the differences observed in the categorisation of the anxiety disorders, the lower proportion of single-item diagnoses of the anxiety disorders (aside from GAD) could be due to a lack of treatment-seeking or recognition. Many individuals with symptoms do not seek treatment for mental health or related problems (27,40) and those that do more commonly discuss their problems with a GP rather than a mental health professional (27). However, research has shown that there is an under-recognition of anxiety disorders, particularly by GPs (50–53). GPs have limited amounts of time and resources and lack specialised training to conduct comprehensive assessments of anxiety symptoms (54). It is therefore possible that GPs encountering distressed patients may identify symptoms as “anxiety” without specifying a disorder. Notably, in the GLAD and COPING studies, the phrasing of the single-item GAD question encapsulates general nerves or anxiety to account for this, which may have resulted in an overestimate of the number of participants given a GAD diagnosis.

Our finding that single-item panic attacks had higher agreement than single-item panic disorder to algorithm-based panic disorder could be further indication of this under-recognition. Although panic attacks are not specific to panic disorder, they are more recognisable and simpler to diagnose than panic disorder. However, studies have found that the majority of individuals that experience a panic attack do not have panic disorder (35). Consequently, this finding could instead indicate a lack of specificity of the algorithm-based panic disorder measure.

### Conclusion

Large-scale research projects that lack the resources to conduct ‘gold standard’ clinical interviewing commonly utilise questionnaires applying algorithm-based or single-item diagnostic methods. We compared these two approaches and found good comparability between algorithm-based and single-item MDD and “any anxiety” disorder for categorisation of participants with a diagnosis, although performance varied between the case and general population cohorts in this study. Ascertainment of participants with diagnoses for the individual anxiety disorders was largely different depending on which phenotyping measure was applied. Our results suggested that single-item diagnoses may be sufficient depending on the aims of the research and the population under study, but may not be suitable for case-control studies investigating disorder-specific risk factors or outcomes. Notably, prior research provides little insight regarding the validity of single-item or algorithm-based diagnoses against clinical interviews. The differences observed in this study highlighted the need for further validation of these diagnoses against clinical interviews to advise measure selection and ensure translatability of research incorporating these measures.

## Data Availability

Code availability
All data cleaning and analyses were conducted using R version 3.5.3 (32), the tidyverse (33), and caret (34) packages. The full code for the diagnostic algorithms and analyses included in this paper are available at https://github.com/mollyrdavies/GLAD-Diagnostic-algorithms.
Data availability
The data that support the findings of this study are available on request from the corresponding author, TCE. The data are not publicly available due to restrictions outlined in the study protocol and specified to participants during the consent process.

## Acknowledgements

We thank the GLAD Study and NIHR BioResource volunteers for their participation, and gratefully acknowledge the NIHR BioResource centres, NHS Trusts and staff for their contribution. We thank the National Institute for Health Research, NHS Blood and Transplant, and Health Data Research UK as part of the Digital Innovation Hub Programme. This study presents independent research funded by the NIHR Biomedical Research Centre at South London and Maudsley NHS Foundation Trust and King’s College London. Further information can be found at http://brc.slam.nhs.uk/about/core-facilities/bioresource. The views expressed are those of the authors and not necessarily those of the NHS, NIHR, HSC R&D Division, Department of Health and Social Care.

## Abbreviations

CIDI: (Composite International Diagnostic Interview)
CIDI-SF: (Composite International Diagnostic Interview - short form)
SCID: (Structured Clinical Interview for DSM-5)
MDD: (major depressive disorder)
GAD: (generalised anxiety disorder)
DSM-5: (Diagnostic Statistical Manual 5)
NIHR: (National Institute for Health Research)
GLAD: (Genetic Links to Anxiety and Depression)
COPING: (COVID-19 Psychiatry and Neurological Genetics)
NBR: (National Institute for Health Research BioResource)
EHR: (electronic health records)
GP: (general practitioner)

