## Supplemental Materials for "Comparison of algorithm-based versus single-item diagnostic measures of anxiety and depression disorders in the GLAD and COPING cohorts"

Supplementary Materials

Comparison of algorithm-based versus single-item phenotyping measures of anxiety and depression disorders in the GLAD Study cohort (Davies et al, in prep).

### **Appendix 1: Algorithm-based Diagnoses**

#### 1a. Overview

Scoring for Depression and Anxiety Modules (DSM-5)

Modules for the GLAD Study sign-up questionnaire include questionnaires adapted from the CIDI-SF (MDD and GAD) and the DSM-5 (anxiety subtypes) by the Australian Genetics of Depression Study team. We utilised responses to these questionnaires to provide a probable diagnosis for major depressive disorder and the specific anxiety disorders. This document outlines how that probable diagnosis is assessed within the GLAD questionnaire and how each question/response matches to a DSM-5 criterion for the disorder.

As an example:

| Do you have (or have you ever had) a strong fear of any of the following things: Animals (e.g. snakes, birds, rats, insects, dogs, or other animals)  **Yes Is Selected** | **Criterion A**  Marked fear or anxiety about a specific object or situation (e.g., flying, heights, animals, receiving an injection, seeing blood). |
| --- | --- |

On the left is the question presented in the questionnaire; in **bold text** is the qualifying response(s) from participants. On the right is the corresponding DSM-5 criterion required for diagnosis of that disorder. Participants who meet all criteria for the disorder presented below will be scored as having a probable diagnosis of the disorder.

For all disorders, we were unable to account for disturbances that are better explained by other disorders/conditions (a DSM-5 criterion included for all disorders), as this is not within the scope of an online questionnaire.

These diagnoses should not be interpreted as final.

#### 1b. Major depressive disorder

##### DSM-5 Diagnostic criteria:

A. Five (or more) of the following symptoms have been present during the same 2-week period and represent a change from previous functioning: at least one of the symptoms is either (1) depressed mood or (2) loss of interest or pleasure.

Note: Do not include symptoms that are clearly attributable to another medical condition.

1. Depressed mood most of the day, nearly every day, as indicated by either subjective report (e.g., feels sad, empty, hopeless) or observation made by others (e.g., appears tearful). (Note: In children and adolescents, can be irritable mood.)
2. Markedly diminished interest or pleasure in all, or almost all, activities most of the day, nearly every day (as indicated by either subjective account or observation).
3. Significant weight loss when not dieting or weight gain (e.g., a change of more than 5% of body weight in a month) or decrease or increase in appetite nearly every day. (Note: In children, consider failure to make expected weight gain.)
4. Insomnia or hypersomnia nearly every day.
5. Psychomotor agitation or retardation nearly every day (observable by others, not merely subjective feelings of restlessness or being slowed down).
6. Fatigue or loss of energy nearly every day.
7. Feelings of worthlessness or excessive or inappropriate guilt (which may be delusional) nearly every day (not merely self-reproach or guilt about being sick).
8. Diminished ability to think or concentrate, or indecisiveness, nearly every day (either by subjective account or as observed by others).
9. Recurrent thoughts of death (not just fear of dying), recurrent suicidal ideation without a specific plan, or a suicide attempt or a specific plan for committing suicide

B. The symptoms cause clinically significant distress or impairment in social, occupational, or other important areas of functioning.

C. The episode is not attributable to the physiological effects of a substance or to another medical condition.

Note: Criteria A-C represent a major depressive episode.

D. The occurrence of the major depressive episode is not better explained by schizoaffective disorder, schizophrenia, schizophreniform disorder, delusional disorder, or other specified and unspecified schizophrenia spectrum and other psychotic disorders.

E. There has never been a manic episode or a hypomanic episode.

Note: This exclusion does not apply if all of the manic-like or hypomanic-like episodes are substance-induced or are attributable to the physiological effects of another medical condition.

##### Scoring algorithm

**IF (ONE OR MORE OF THE FOLLOWING)**

| Have you ever had a time in your life when you have felt sad, blue, or depressed for two weeks or more in a row?  **Yes Is Selected** | **Criterion A**  **Depressed mood** most of the day, nearly every day for a **2-week period**, as indicated by either subjective report or observation made by others. |
| --- | --- |
| *Variable name:*  **cidid.low_mood –** Yes (1) |  |

*AND/OR*

| Have you ever had a time in your life lasting two weeks or more when you lost interest in most things like hobbies, work, or activities that usually give you pleasure?  **Yes is Selected** | **Criterion A**  **Markedly diminished interest or pleasure in all, or almost all, activities** most of the day, nearly every day for a **2-week period**, as indicated by either subjective account or observation. |
| --- | --- |
| *Variable name:*  **cidid.anhedonia** **–** Yes (1) | |

**AND**

| How much of the day did these feelings usually last?   - All day long **Is Selected**   **OR**   - Most of the day **Is Selected** | **Criterion A**  Depressed mood/markedly diminished interest in activities **most of the day,** nearly every day, as indicated by either subjective report or observation. |
| --- | --- |
| *Variable name:*  **cidid.how_long** **–** All day long (4) **OR** Most of the day (3) | |

**AND**

| How often did you feel this way?   - Every day **Is Selected**   **OR**   - Almost every day **Is Selected** | **Criterion A**  Depressed mood/markedly diminished interest in activities most of the day**, nearly every day**, as indicated by either subjective report or observation. |
| --- | --- |
| *Variable name:*  **cidid.how_often** – Every day (3) OR Almost every day (2) | |

**AND (FIVE OR MORE OF THE FOLLOWING)**

| Have you ever had a time in your life when you have felt sad, blue, or depressed for two weeks or more in a row?  **Yes Is Selected** | **Criterion A**  Depressed mood most of the day, nearly every day, as indicated by either subjective report or observation made by others. |
| --- | --- |
| *Variable name:*  **cidid.low_mood** – Yes (1) | |

*Note: this criterion is also mentioned above because it’s one of the two cardinal symptoms, but is also included in the 5+ symptoms required for a diagnosis*

*AND/OR*

| Have you ever had a time in your life lasting two weeks or more when you lost interest in most things like hobbies, work, or activities that usually give you pleasure?  **Yes is Selected** | **Criterion A**  Markedly diminished interest or pleasure in all, or almost all, activities most of the day, nearly every day (as indicated by either subjective account or observation). |
| --- | --- |
| *Variable name:*  **cidid.anhedonia** – Yes (1) | |

*Note: this criterion is also mentioned above because it’s one of the two cardinal symptoms, but is also included in the 5+ symptoms required for a diagnosis*

*AND/OR*

| Did your weight change? | **Criterion A**  Significant weight loss when not dieting or weight gain (e.g., a change of more than 5% of body weight in a month), or decrease or increase in appetite nearly every day. (Note: In children, consider failure to make expected weight gain.) |
| --- | --- |
| - Gained weight **Is Selected**   **OR** |  |
| - Lost weight **Is Selected**   **OR**   - Both gained and lost some weight during the episode **Is Selected**   AND (optional, not currently included)  Did your weight change by about 10lbs (4kg) or more?  **Yes Is Selected**  *AND/OR*  Did you experience a change in your appetite?   - Increased appetite **Is Selected**   **OR**   - Decreased appetite **Is Selected**   **OR**   - Experienced both increased and decreased appetite during the same depression episode **Is Selected** |  |
| *Variable name:*  **cidid.weight_change** – Gained weight (1) **OR** Lost weight (2) **OR** Both (3)  *AND/OR* **cidid.appetite_change –** Increased appetite (1) **OR** Decreased appetite (2) **OR** Both increased and decreased appetite (3) | |

*AND/OR*

| Did your sleep change?  **Yes Is selected** | **Criterion A**  Insomnia or hypersomnia nearly every day. |
| --- | --- |
| *Variable name:*  **cidid.sleep_change** – Yes (1) | |

*AND/OR*

| [NOT MEASURED]  .. **Is Selected** | **Criterion A**  Psychomotor agitation or retardation nearly every day (observable by others, not merely subjective feelings of restlessness or being slowed down). |
| --- | --- |

*AND/OR*

| Did you feel more tired out or low on energy than is usual for you?  **Yes Is Selected** | **Criterion A**  Fatigue or loss of energy nearly every day. |
| --- | --- |
| *Variable name:*  **cidid.fatigue** – Yes (1) | |

*AND/OR*

| People sometimes feel down on themselves, no good, worthless. Did you feel this way?  **Yes Is Selected** | **Criterion A**  Feelings of worthlessness or excessive or inappropriate guilt (which may be delusional) nearly every day (not merely self-reproach or guilt about being sick). |
| --- | --- |
| *Variable name:*  **cidid.worthlessness** – Yes (1) | |

*AND/OR*

| Did you have a lot more trouble concentrating than usual?  **Yes Is Selected** | **Criterion A**  Diminished ability to think or concentrate, or indecisiveness, nearly every day (either by subjective account or as observed by others). |
| --- | --- |
| *Variable name:*  **cidid.trouble_concentrating** – Yes (1) | |

*AND/OR*

| Did you think a lot about death - either your own, someone else's, or death in general?  **Yes Is Selected** | **Criterion A**  Recurrent thoughts of death (not just fear of dying), recurrent suicidal ideation without a specific plan, or a suicide attempt or a specific plan for committing suicide. |
| --- | --- |
| *Variable name:*  **cidid.thoughts_of_death** – Yes (1) | |

**AND**

| Think about your roles at the time of this episode, including study/employment, childcare and housework, leisure pursuits. How much did these problems interfere with your life or activities?   - **A lot Is Selected**   **OR**   - **Some Is Selected** | **Criterion B**  The symptoms cause clinically significant distress or impairment in social, occupational, or other important areas of functioning. |
| --- | --- |
| *Variable name:*  **cidid.functioning** – A lot (3) or Some (2) | |

*(The options for this question include: ‘a lot,’ ‘some,’ ‘a little’ or ‘none.’ We have drawn the line of clinically significant distress at ‘some’ or ‘a lot.’)*

##### CIDI-SF measure

*Note that the CIDI-SF measure is a portion of the MDD section of the questionnaire, therefore the numbering does not start at 1.*

**B2) Have you ever had a time in your life when you felt sad, blue, or depressed for two weeks or more in a row?**

🞏 Yes

🞏 No

🞏 Prefer not to answer

**B3) Have you ever had a time in your life lasting two weeks or more when you lost interest in most things like hobbies, work, or activities that usually give you pleasure?**

🞏 Yes

🞏 No

🞏 Prefer not to answer

**If ‘Yes’ to either B2 or B3, continue to B4. If ‘No’/‘Prefer not to answer’ to both B2 and B3, skip to B27 (Mood Disorder Questionnaire)**

**B4)** **Please think of the two-week period in your life when your feelings of depression or loss of interest were worst:**

**B4a) How much of the day did these feelings usually last?**

| 🞏 All day long | 🞏 Most of the day | 🞏 About half of the day |
| --- | --- | --- |
| 🞏 Less than half of the day | 🞏 Don’t know | 🞏 Prefer not to answer |

**B4b) Did you feel this way:**

| 🞏 Every day | 🞏 Almost every day | 🞏 Less often |
| --- | --- | --- |
| 🞏 Don’t know | 🞏 Prefer not to answer |  |

**B5) Did you feel more tired out or low on energy than is usual for you?**

| 🞏 Yes | 🞏 No |
| --- | --- |
| 🞏 Don’t know | 🞏 Prefer not to answer |

**In the next section (B6-B19), we would like to know more about your depression or low mood. *If you find this is too difficult, you can skip to B20 on pg.5.***

**B6) Did your weight change? (***do not include weight change as a side-effect of medication you were taking***)**

| 🞏 Gained weight *(continue to B6a)* | 🞏 Lost weight *(continue to B6a)* | 🞏 Both gained weight and lost some  weight during the episode (*continue to B6a*) |
| --- | --- | --- |
| 🞏 Stayed about the same or was on  a diet *(skip to B7)* | 🞏 Don’t know *(skip to B7)* | 🞏 Prefer not to answer *(skip to B7)* |

**B6a) Did your weight change by about 10lbs (4kg) or more?**

| 🞏 Yes | 🞏 No |
| --- | --- |
| 🞏 Don’t know | 🞏 Prefer not to answer |

**B7) Did your sleep change? (***do not include sleep change as a side-effect of medication you were taking***)**

🞏 Yes *(continue to B7a)*

🞏 No *(skip to B8)*

🞏 Don’t know *(skip to B8)*

🞏 Prefer not to answer *(skip to B8)*

| **B7a) Was that:** *(please select all that apply)* | **1**  **Yes** |  | **2**  **No** |
| --- | --- | --- | --- |
| Trouble falling asleep (sleeping too little) | 🞏 |  | 🞏 |
| Waking too early (sleeping too little) | 🞏 |  | 🞏 |
| Sleeping too much | 🞏 |  | 🞏 |
| Both sleeping too much and too little during the same depression episode | 🞏 |  | 🞏 |

**Any ‘Yes’ selections made in B7a lead to display of B7b and B7c**

**B7b)** **How many hours per day did you sleep on average during the depression episode, including nighttime sleep and daytime naps?**

🞏🞏

**B7c)** **How many hours per day did you used to sleep on average when you were not depressed?**

🞏🞏

**B8) Did you experience a change in your appetite?**

| 🞏 No changes in appetite | 🞏 Increased appetite |
| --- | --- |
| 🞏 Decreased appetite | 🞏Experienced both increased an decreased appetite during the same depression episode |
| 🞏Don’t know | 🞏Prefer not to answer |

**B9) Did your mood brighten in response to positive events?**

| 🞏 Yes | 🞏 No |
| --- | --- |
| 🞏 Don’t know | 🞏 Prefer not to answer |

**B10) Did you experience heavy feelings in your arms or legs? (Did your arms or legs feel “heavy”?)**

| 🞏 Yes *(continue to B10.1)* | 🞏 No *(skip to B11)* |
| --- | --- |
| 🞏 Don’t know *(skip to B11)* | 🞏 Prefer not to answer *(skip to B11)* |

**B10.1 For how many hours per day was the heaviness feeling present?**

🞏🞏

**B11) Where you overly sensitive to interpersonal rejection?**

🞏 No

🞏 Yes, and this significantly impaired your social or work relationships

🞏 Yes, but this did not significantly impair your social or work relationships

🞏 Don’t know

🞏 Prefer not to answer

**B12) Was your mood worse:**

🞏 In the morning

🞏 In the afternoon

🞏 At night

🞏 My mood did not vary

🞏 Don’t know

🞏 Prefer not to answer

|  | **1**  **Yes** | | **2**  **No** | | | **3**  **Don’t Know** | **4**  **Prefer not to answer** | |
| --- | --- | --- | --- | --- | --- | --- | --- | --- |
| **B13)** Did you have a lot more trouble concentrating than usual? | | 🞏 | | 🞏 | 🞏 | | | 🞏 |
| **B14)** People sometimes feel down on themselves, no good, worthless. Did you feel this way? | | 🞏 | | 🞏 | 🞏 | | | 🞏 |
| **B15)** Did you think a lot about death – either your own, someone else’s or death in general? | | 🞏 | | 🞏 | 🞏 | | | 🞏 |

**B16) Roughly how long altogether did you feel this way?**

| 🞏 Less than a month | 🞏 Between one  and three months | 🞏 Over three months, but  less than six months | 🞏 Over six months,  but less than 12 months |
| --- | --- | --- | --- |
| 🞏 One to two years | 🞏Over two years | 🞏Don’t know | 🞏Prefer not to answer |

**B17 Was this your longest episode of depression or low mood?**

| 🞏 Yes (skip to B18) | 🞏 No | 🞏 Don’t know (skip to B18) | 🞏 Prefer not to answer (skip to B18) |
| --- | --- | --- | --- |

**B17a) What is the longest period of time that you have experienced depression or low mood?**

| 🞏 Less than 6 months | 🞏 Between 6 and 12 months |
| --- | --- |
| 🞏 Between 1 and 5 years | 🞏 More than 5 years |
| 🞏All of my life / as long as I can remember |  |

**B18) Think about your roles at the time of this episode, including study/employment, childcare and housework, leisure pursuits. How much did these problems interfere with your life or activities?**

| 🞏 A lot | 🞏 Some | 🞏 A little |
| --- | --- | --- |
| 🞏 Not at all | 🞏Prefer not to answer |  |

**B19) How many periods of depression or low mood have you had in your life lasting two or more weeks?**

| 🞏 One *(skip to B20)* | 🞏Two – three *(continue to B19a)* |
| --- | --- |
| 🞏 Several *(continue to B19a)* | 🞏All of my life / as long as I can remember *(continue to B20)*  🞏 Prefer not to answer *(skip to B20)* |

**B19a) If you have had several, please estimate the number of times you have had periods of depression or low mood in your life lasting two or more weeks:**

🞏🞏

**B20) About how old were you the first time you had a period of two weeks like this? (Whether or not you received any help for it)**

🞏🞏

**B21 is not displayed to those who answered “One” in B19 or B19a**

**B21) About how old were you the last time you had a period of two weeks like this? (Whether or not you received any help for it)**

🞏🞏

**B22 is only displayed for participants indicating Yes to Q3 “Have you ever been pregnant?”**

**B22) Did any of these episodes occur within the first year of giving birth? Or has it been suggested you had post-natal depression?**

| 🞏 Yes | 🞏 No | 🞏 Don’t know |
| --- | --- | --- |
| 🞏 Prefer not to answer | 🞏 Not applicable |  |

**B22.1) Did any of these episodes occur following a significant or traumatic event such as death/serious illness of close relative or friend, or following an distressing event or illness that happened to you?**

| 🞏 Most/All | 🞏 More than once | 🞏 Once |
| --- | --- | --- |
| 🞏 Not at all | 🞏Prefer not to answer |  |

**B23) Did you ever tell a professional about these problems?** *(Medical doctor, psychologist, social worker, counsellor, nurse, clergy, or other helping professional)*

| 🞏 Yes | 🞏 No |
| --- | --- |
| 🞏 Don’t know | 🞏 Prefer not to answer |

**B24) Did you ever try the following for these problems?**

Tick ALL that apply:

| 🞏 Medication prescribed to  you for at least two weeks | 🞏 Unprescribed medication  more than once | 🞏 Drugs or alcohol more than once |
| --- | --- | --- |
| 🞏Psychotherapy or other talking therapy more than once (including internet-based CBT)  🞏Regular physical exercise (e.g. yoga, running, walking) | 🞏Structured wellbeing activity (e.g. mindfulness, meditation, self-help book) | 🞏 Prefer not to answer  🞏None of the above |

**B24a is only displayed to those who selected ‘psychotherapy of other talking therapy’ in B24**

**B24a) Are you currently enrolled in an NHS funded talking therapy or psychotherapy (IAPT)?**

| 🞏 Yes | 🞏 No |
| --- | --- |
| 🞏 Don’t know |  |

**B24b and B24c are only displayed to those who selected ‘prescribed medication’ in B23**

**B24b) Did you take your medication as advised?**

| 🞏 Yes | 🞏 No |
| --- | --- |
| 🞏 Don’t know | 🞏 Prefer not to answer |

**B24c) Did you find the medication helpful?**

| 🞏 Yes | 🞏 No |
| --- | --- |
| 🞏 Don’t know | 🞏 Prefer not to answer |

**B25 – B25b is only displayed to those who selected ‘psychotherapy or other talking therapy’ or ‘structured wellbeing activity’ in B24**

**B25) You previously mentioned that you have tried psychotherapy or another talking therapy, please select all that you attended more than once.**

| 🞏 Counselling | 🞏 Group therapy | 🞏 Cognitive Behavioral Therapy (CBT) |
| --- | --- | --- |
| 🞏 Mindfulness | 🞏 Guided self-help | 🞏 Workshops |
| 🞏 Relationship therapy | 🞏 Family therapy | 🞏 Online therapy |
| 🞏 Never tried psychotherapy or other talking therapies | 🞏 Prefer not to answer | 🞏 Other (*please specify:* _____________________) |

**25a) Did you complete your course of psychotherapy or other talking therapy?**

| 🞏 Yes | 🞏 No |
| --- | --- |
| 🞏 Don’t know | 🞏 Prefer not to answer |

**25b) Did you find psychotherapy or other talking therapy helpful?**

| 🞏 Yes | 🞏 No |
| --- | --- |
| 🞏 Don’t know | 🞏 Prefer not to answer |

**Questions B26.1-B26.4 will only display to participants with a score of 5 or more on B1 and selected ‘yes’ to either B2 or ‘yes’ to B3**

**B26.1) How long ago did your current or most recent episode of depression or low mood begin?**

🞏 Less than 1 year ago *(continue to B1.2)*

🞏 1 to 2 years ago *(continue to B1.2)*

🞏 More than 2 years ago *(continue to B1.2)*

**B26.2) During the current or most recent episode of depression or low mood, how many antidepressant medications have you taken for 6 weeks or longer?**

🞏 I have not taken medication during my current episode of depression *(continue to B1.3)*

🞏 1-2 medications *(continue to B1.3)*

🞏 3-4 medications *(continue to B1.3)*

🞏 5-6 medications *(continue to B1.3)*

🞏 7-10 medications *(continue to B1.3)*

🞏 >10 medications *(continue to B1.3)*

**B26.3) If people don’t respond fully to antidepressants, doctors sometime prescribe “add-on” or “augmentation” medications in addition to the antidepressant (such as lithium, quetiapine or aripiprazole).

During the current or most recent episode of depression or low mood, have you taken an add-on medication for 6 weeks or longer?**

🞏 Yes *(continue to B1.4)*

🞏 No *(continue to B1.4)*

🞏 Prefer not to answer *(continue to B1.4)*

**B26.4) Have you received electroconvulsive therapy (ECT) in this current or most recent episode of depression or low mood? (Please only answer yes if your course of ECT included 8 treatment sessions or more.)**

🞏 Yes *(continue to B2*

🞏 No *(continue to B2)*

🞏 Prefer not to answer *(continue to B2)*

#### 1c. Generalised anxiety disorder

##### DSM-5 Diagnostic criteria

A. Excessive anxiety and worry (apprehensive expectation), occurring more days than not for at least 6 months, about a number of events or activities (such as work or school performance).

B. The individual finds it difficult to control the worry.

C. The anxiety and worry are associated with three (or more) of the following six symptoms (with at least some symptoms having been present for more days than not for the past 6 months)

1. Restlessness or feeling keyed up or on edge.
2. Being easily fatigued.
3. Difficulty concentrating or mind going blank.
4. Irritability.
5. Muscle tension.
6. Sleep disturbance (difficulty falling or staying asleep, or restless, unsatisfying sleep).

D. The anxiety, worry, or physical symptoms cause clinically significant distress or impairment in social, occupational, or other important areas of functioning.

E. The disturbance is not attributable to the physiological effects of a substance (e.g., a drug of abuse, a medication) or another medical condition (e.g., hyperthyroidism).

F. The disturbance is not better explained by another mental disorder

##### Scoring algorithm

**IF (ONE OR MORE OF THE FOLLOWING)**

| Have you ever had a period lasting one month or longer when most of the time you felt worried, tense, or anxious?  **Yes Is Selected**  **AND/OR**  People differ a lot in how much they worry about things. Did you ever have a time when you worried a lot more than most people would in your situation?  **Yes Is Selected** | **Criterion A**  Excessive anxiety and worry occurring more days than not for at least 6 months, about a number of events or activities. |
| --- | --- |
| *Variable name:*  **cidia.felt_worried** – Yes (1) **AND/OR** **cidia.felt_worried_more** – Yes (1) | |

*(Note: There is another variable in the questionnaire that asks: “During that period, was your worry stronger than in other people?” (variable name: cidia.worry_stronger_than_others). It is not included in the algorithm because the other two variables are screening variables. Participants are not shown the remaining questions if they respond ‘No’ to both, so this additional variable has no utility.)*

**AND**

| What is the longest period of time that this kind of worrying has ever continued?   - Between 6 and 12 months **Is Selected**   **OR**   - Between 1 and 5 years **Is Selected**   **OR**   - More than 5 years **Is Selected**   **OR**   - All my life / as long as I can remember **Is Selected** | **Criterion A**  Excessive anxiety and worry occurring more days than not for at least 6 months, about a number of events or activities. |
| --- | --- |
| *Variable name:*  **cidia.longest_period_worry_categorical*** – Between 6 and 12 months (2) **OR** Between 1 and 5 years (3) **OR** More than 5 years (4) **OR** All my life / as long as I can remember (5) | |

*Note: at the start of GLAD, the longest period variable consisted of 2 free text fields that needed to be converted to assess >6 months duration. Variable name: cidia.longest_period_worry_years & cidia. longest_period_worry_months]

**AND**

| Did you worry most days?  **Yes Is Selected** | **Criterion A**  Excessive anxiety and worry occurring **more days than not** for at least 6 months, about a number of events or activities. |
| --- | --- |
| *Variable name:*  **cidia.most_days** – Yes (1) | |

**AND**

| Did you usually worry about one particular thing, such as your job security or the failing health of a loved one, or more than one thing?  **More than one thing Is Selected**  **AND/OR**  Did you ever have different worries on your mind at the same time?  **Yes Is Selected** | **Criterion A**  Excessive anxiety and worry occurring more days than not for at least 6 months, about a **number of events or activities.** |
| --- | --- |
| *Variable names:*  **cidia.more_than_one_thing** – More than one thing (1) **AND/OR cidia.different_worries** – Yes (1) | |

**AND**

| Did you find it difficult to stop worrying?  **Yes Is Selected**  **AND/OR**  How often was your worry so strong that you couldn't put it out of your mind no matter how hard you tried?  **Often Is selected**  **AND/OR**  How often did you find it difficult to control your worry?  **Often Is selected** | **Criterion B**  The person finds it difficult to control the worry. |
| --- | --- |
| *Variable names:*  **cidia.difficult_to_stop –** Yes (1) **AND/OR cidia.couldnt_stop** – Often (3) **AND/OR cidia.difficult_to_control –** Often (3) | |

**AND (THREE OR MORE OF THE FOLLOWING)**

| Restless?  **Yes Is Selected**  **AND/OR**  Keyed up or on edge?  **Yes Is Selected** | **Criterion C**  Restlessness or feeling keyed up or on edge |
| --- | --- |
| *Variable names:*  **cidia.restless AND/OR cidia.on_edge** – Yes (1) | |

*AND/OR*

| Easily tired?  **Yes Is selected** | **Criterion C**  Being easily fatigued |
| --- | --- |
| *Variable name:*  **cidia.tired** – Yes (1) | |

*AND/OR*

| Having difficulty keeping your mind on what you were doing?  **Yes Is selected** | **Criterion C**  Difficulty concentrating or mind going blank |
| --- | --- |
| *Variable name:*  **cidia.difficulty_concentrating** – Yes (1) | |

*AND/OR*

| More irritable than usual?  **Yes Is Selected** | **Criterion C**  Irritabiity |
| --- | --- |
| *Variable name:*  **cidia.irritable** – Yes (1) | |

*AND/OR*

| Having tense, sore, or aching muscles?  **Yes Is Selected** | **Criterion C**  Muscle tension |
| --- | --- |
| *Variable name:*  **cidia.tense_muscles** – Yes (1) | |

*AND/OR*

| Often having trouble falling or staying asleep?  **Yes Is Selected** | **Criterion C**  Sleep disturbance (difficulty falling or staying asleep, or restless unsatisfying sleep) |
| --- | --- |
| *Variable name:*  **cidia.trouble_sleeping** – Yes (1) | |

**AND**

| Regarding times in your life when you have felt worried, tense or anxious:  Think about your roles at the time of this episode, including study/employment, childcare and housework, leisure pursuits. How much did these problems interfere with your life or activities?  **Some Is Selected**  **OR**  **A lot is selected** | **Criterion E**  The anxiety, worry, or physical symptoms cause clinically significant distress or impairment in social, occupational, or other important areas of functioning. |
| --- | --- |
| *Variable name:*  **cidia.functioning –** Some (2) **OR** A lot (3) | |

*(The options for this question include: ‘a lot,’ ‘some,’ ‘a little’ or ‘none.’ We have drawn the line of clinically significant distress at ‘some’ or ‘a lot.’)*

##### CIDI-SF measure

*Note that the CIDI-SF measure is a portion of the GAD section of the questionnaire, therefore the numbering does not start at 1.*

**C2a) Have you ever had a period lasting one month or longer when most of the time you felt worried, tense, or anxious?**

🞏 Yes

🞏 No

🞏 Don’t know

🞏 Prefer not to answer

**C2b) People differ a lot in how much they worry about things. Did you ever have a time when you worried a lot more than most people would in your situation?**

🞏 Yes

🞏 No

🞏 Don’t know

🞏 Prefer not to answer

**Participants who answer ‘No’ or ‘Prefer not to answer’ on C2a and C2b skip to Section D.**

**C2c) What is the longest period of time that this kind of worrying has ever continued?**

| 🞏 Less than 6 months *(skip to Section D)* | 🞏 Between 6 and 12 months |
| --- | --- |
| 🞏 Between 1 and 5 years | 🞏 More than 5 years |
| 🞏All of my life / as long as I can remember |  |

**C3a) How many periods of this kind of worry have you had in your life lasting 6 or more months?**

| 🞏 One *(skip to C3c)* | 🞏 Two-three | |
| --- | --- | --- |
| 🞏 Several | | 🞏All of my life / as long as I can remember |
| 🞏Prefer not to answer *(skip to C3c)* |  | |

**C3b) Please estimate the number of times you have had periods of this kind of worry in your life lasting 6 or more months:**

🞏🞏

**C3c) About how old were you the first time you had a period of six months like this? (Whether or not you received any help for it.) Please put your age in years. An approximate age is fine.**

🞏🞏

**C3d is not displayed to those who answered “one” in C2b or in C2c**

**C3d) About how old were you the last time you had a period of six months like this? (Whether or not you received any help for it.) Please put your age in years. An approximate age is fine.**

🞏🞏

**Please think of the period in your life when you have felt worried, tense, anxious, or more worried than most people would in your situation. This could be in the past, or it could be continuing now.**

Tick an answer for each statement:

|  | **1**  **Yes** | **2**  **No** | **3**  **Don’t Know** | **4**  **Prefer not to answer** |
| --- | --- | --- | --- | --- |
| **C4) During that period, was your worry stronger than in other people?** | 🞏 | 🞏 | 🞏 | 🞏 |
| **C5) Did you worry most days?** | 🞏 | 🞏 | 🞏 | 🞏 |

|  | **1**  **One thing** | **2**  **More than one thing** | **3**  **Don’t Know** | **4**  **Prefer not to answer** |
| --- | --- | --- | --- | --- |
| **C6) Did you usually worry about one particular thing, such as your job security or the failing health of a loved one, or more than one thing?** | 🞏 | 🞏 | 🞏 | 🞏 |

|  | **1**  **Yes** | **2**  **No** | **3**  **Don’t Know** | **4**  **Prefer not to answer** |
| --- | --- | --- | --- | --- |
| **C7) Did you find it difficult to stop worrying?** | 🞏 | 🞏 | 🞏 | 🞏 |
| **C8) Did you ever have different worries on your mind at the same time?** | 🞏 | 🞏 | 🞏 | 🞏 |

|  | **1**  **Often** | **2**  **Sometimes** | **3**  **Rarely** | **4**  **Never** | **5**  **Don’t know** | **6**  **Prefer not to say** |
| --- | --- | --- | --- | --- | --- | --- |
| **C9) How often was your worry so strong that you couldn’t put it out of your mind no matter how hard you tried?** | 🞏 | 🞏 | 🞏 | 🞏 | 🞏 | 🞏 |
| **C10) How often did you find it difficult to control your worry?** | 🞏 | 🞏 | 🞏 | 🞏 | 🞏 | 🞏 |

**C11) When you were worried or anxious, were you also:**

Tick an answer for each statement:

|  | **1**  **Yes** | **2**  **No** | **3**  **Don’t Know** |
| --- | --- | --- | --- |
| **a.** Restless? | 🞏 | 🞏 | 🞏 |
| **b.** Keyed up or on edge? | 🞏 | 🞏 | 🞏 |
| **c.** Easily tired? | 🞏 | 🞏 | 🞏 |
| **d.** Having difficulty keeping your mind on what you were doing? | 🞏 | 🞏 | 🞏 |
| **e.** More irritable than usual? | 🞏 | 🞏 | 🞏 |
| **f.** Having tense, sore, or aching muscles? | 🞏 | 🞏 | 🞏 |
| **g.** Often have trouble falling or staying asleep? | 🞏 | 🞏 | 🞏 |

**C12) Did you ever tell a professional about these problems (medical doctor, psychologist, social worker, counsellor, nurse, clergy, or other helping professional)?**

| 🞏 Yes | 🞏 No |
| --- | --- |
| 🞏 Don’t know | 🞏 Prefer not to answer |

**C13) Did you ever use the following for the worry or the problems it caused? Please include any treatments that you have already told us about under ‘depression’ if they were also for anxiety:**

**Tick ALL that apply**

| 🞏 Medication prescribed to  you for at least two weeks | 🞏 Specific anti-anxiety medication prescribed to you for at least one week | 🞏 Unprescribed medication  more than once |
| --- | --- | --- |
| 🞏 Drugs or alcohol more than once | 🞏Psychotherapy or other talking therapy more than once (including internet-based CBT) | 🞏Structured wellbeing activity (e.g. mindfulness, meditation, self-help book) |
| 🞏Regular physical exercise (e.g. yoga, running, walking) | 🞏 Prefer not to answer | 🞏None of the above |

**C13a is only displayed to those who selected ‘psychotherapy or other talking therapy’ in C13**

**C13a) Are you currently enrolled in an NHS funded talking therapy or psychotherapy (IAPT)?**

| 🞏 Yes | 🞏 No |
| --- | --- |
| 🞏 Don’t know |  |

**C13b and C13c is only displayed to those who selected ‘prescribed medication’ in C13**

**C13b) Did you take your medication as advised?**

| 🞏 Yes | 🞏 No |
| --- | --- |
| 🞏 Don’t know | 🞏 Prefer not to answer |

**C13cb) Did you find the medication helpful?**

| 🞏 Yes | 🞏 No |
| --- | --- |
| 🞏 Don’t know | 🞏 Prefer not to answer |

**C14-C14b are only displayed to those who selected ‘psychotherapy or other talking therapy’ or ‘structured wellbeing activity’ in C13**

**C14) You previously mentioned that you have tried psychotherapy or another talking therapy, please select all that you attended more than once.**

| 🞏 Counselling | 🞏 Group therapy | 🞏 Cognitive Behavioral Therapy (CBT) |
| --- | --- | --- |
| 🞏 Psychotherapy | 🞏 Mindfulness | 🞏 Guided self-help |
| 🞏 Workshops | 🞏 Online therapy | 🞏 Psychodynamic |
| 🞏 Psychoanalysis | 🞏 Family therapy | 🞏 Relationship therapy |
| 🞏 Never tried psychotherapy or other talking therapies *(skip to C15)* | 🞏 Prefer not to answer | 🞏 Other |

**C14a) Did you complete your course of psychotherapy or other talking therapy?**

| 🞏 Yes | 🞏 No |
| --- | --- |
| 🞏 Don’t know | 🞏 Prefer not to answer |

**C14b) Did you find psychotherapy or other talking therapy helpful?**

| 🞏 Yes | 🞏 No |
| --- | --- |
| 🞏 Don’t know | 🞏 Prefer not to answer |

**C15) Think about your roles at the time of this episode, including study/employment, childcare and housework, leisure pursuits. How much did these problems interfere with your life or activities?**

| 🞏 A lot | 🞏 Some | 🞏 A little |
| --- | --- | --- |
| 🞏 Not at all | 🞏Prefer not to answer |  |

#### 1d. Specific phobia

##### DSM-5 Diagnostic criteria:

A. Marked fear or anxiety about a specific object or situation (e.g., flying, heights, animals, receiving an injection, seeing blood).

B. The phobic object or situation almost always provokes immediate fear or anxiety.

C. The phobic object or situation is actively avoided or endured with intense fear or anxiety.

D. The fear or anxiety is out of proportion to the actual danger posed by the specific objector situation and to the sociocultural context.

E. The fear, anxiety, or avoidance is persistent, typically lasting for 6 months or more.

F. The fear, anxiety, or avoidance causes clinically significant distress or impairment in social, occupational, or other important areas of functioning.

G. The disturbance is not better explained by the symptoms of another mental disorder, including fear, anxiety, and avoidance of situations associated with panic-like symptoms or other incapacitating symptoms (as in agoraphobia): objects or situations related to obsessions (as in obsessive-compulsive disorder); reminders of traumatic events (as in posttraumatic stress disorder); separation from home or attachment figures (as in sep­aration anxiety disorder); or social situations (as in social anxiety disorder).

##### Scoring algorithm

**IF**

| Do you have (or have you ever had) a strong fear of any of the following things:   - Animals (e.g. snakes, birds, rats, insects, dogs, or other animals)   **Yes Is Selected**  **AND/OR**   - Environment (e.g. heights, storms, thunder, lightning, or being in still water, like a swimming pool or lake)   **Yes Is Selected**  **AND/OR**   - Blood, injections or injury (e.g. blood, needles, medical procedures) -   **Yes Is Selected**  **AND/OR**   - Situations (e.g. being in an airplane, elevator, or a closed space like a cave or tunnel)   **Yes Is Selected**  **AND/OR**   - Other (e.g. situations that may lead to choking or vomiting)   **Yes Is Selected** | **Criterion A**  Marked fear or anxiety about a specific object or situation (e.g., flying, heights, animals, receiving an injection, seeing blood). |
| --- | --- |
| *Variable name:*  **spec.environment_phobia** – Yes (1) **OR** **spec.situation_phobia** – Yes (1) **OR spec.animal_phobia** – Yes (1) **OR** **spec.blood_injection_phobia** – Yes (1) **OR spec.other_phobia** – Yes (1) | |

**AND**

| How often do (or did) these situations cause immediate fear or anxiety for you?   - **Almost always Is Selected**   **OR**   - **Always is selected** | **Criterion B**  The phobic object or situation almost always provokes immediate fear or anxiety. |
| --- | --- |
| *Variable name:*  **spec.phobia_frequency** – Almost always (3) **OR** Always (4) | |

**AND**

| Do you (or did you) | **Criterion C**  The phobic object or situation is actively avoided or endured with intense fear or anxiety. |
| --- | --- |
| - Avoid these situations?   **Yes is selected**  **AND/OR** |  |
| - Endure them with intense anxiety?   **Yes Is selected** |  |
| *Variable name:*  **spec.avoid_phobias** – Yes (1) **OR spec.endure_phobias_with_anxiety** – Yes (1) | |

**AND**

| Are (or were) any of these fears out of proportion to the actual danger involved?  **Yes Is selected** | **Criterion D**  The fear or anxiety is out of proportion to the actual danger posed by the specific object or situation and  to the sociocultural context. |
| --- | --- |
| *Variable name:*  **spec.phobia_out_of_proportion** – Yes (1) | |

**AND**

| How long was the longest time any of these fears lasted?   - Between 6 and 12 months **Is Selected**   **OR**   - Between 1 and 5 years **Is Selected**   **OR**   - More than 5 years **Is Selected**   **OR**   - All my life / as long as I can remember **Is Selected** | **Criterion E**  The fear, anxiety, or avoidance is persistent, typically lasting for 6 months or more. |
| --- | --- |
| *Variable name:*  **spec.phobia_lasted** – 6-12 months (2) **OR** 1-5 years (3) **OR** 5+ years (4) **OR** All of my life (5) | |

**AND**

| How much have any of these fears ever interfered with your life or activities?   - **Some Is Selected**   **OR**   - **A lot is selected** | **Criterion F**  The fear, anxiety, or avoidance causes clinically significant distress or impairment in social, occupational, or other important areas of functioning. |
| --- | --- |
| *Variable name:*  **spec.phobia_interfered –** Some (2) **OR** A lot (3) | |

*(The options for this question include: ‘a lot,’ ‘some,’ ‘a little’ or ‘none.’ We have drawn the line of clinically significant distress at ‘some’ or ‘a lot.’)*

##### CIDI-SF measure

**The next questions are about things that make some people so afraid that they avoid them or they endure them with intense fear or anxiety.**

**1) Do you have (or have you ever had) a strong fear of any of the following things:**

|  | **1**  **No** | **2**  **Yes** |
| --- | --- | --- |
| **a. Environment (e.g. heights, storms, thunder, lightning, or being in still water, like a swimming pool or lake)** | 🞏 | 🞏 |
| **b. Situations (e.g. being in an airplane, elevator, or a closed space like a cave or tunnel)** | 🞏 | 🞏 |
| **c. Animals (e.g. snakes, birds, rats, insects, dogs, or other animals)** | 🞏 | 🞏 |
| **d. Blood, injections or injury (e.g. blood, needles, medical procedures)** | 🞏 | 🞏 |
| **e. Other (e.g. situations that may lead to choking or vomiting)** | 🞏 | 🞏 |

**1.1) Do you have a fear of vomiting?**

| 🞏 Yes | 🞏 No |
| --- | --- |

**If no is selected for all the statements in question 1 please skip to the next section. If yes is selected for one or more of these statements, please continue to question 1.2.**

**1.2) Do you (or did you)…?**

|  | **1**  **No** | **2**  **Yes** |
| --- | --- | --- |
| **a. Avoid these situations?** | 🞏 | 🞏 |
| **b. Endure them with intense anxiety?** | 🞏 | 🞏 |

**2) Thinking about the situations that you fear (or feared)**

**How often do (or did) these situations cause immediate fear or anxiety for you?**

| 🞏 Always | 🞏 Almost always |
| --- | --- |
| 🞏 Some of the time *(skip to next section)* | 🞏 Only one or two times ever *(skip to next section)* |
| 🞏Never *(skip to next section)* |  |

**3) How old were you when these fears first started?**

🞏🞏

**4) How old were you when you most recently experienced these fears?**

🞏🞏

**5) How long was the longest time any of these fears lasted?**

| 🞏 Less than 6 months | 🞏 Between 6 and 12 months |
| --- | --- |
| 🞏 Between 1 and 5 years | 🞏 More than 5 years |
| 🞏All of my life / as long as I can remember |  |

**6) How much have any of these fears ever interfered with your life or activities?**

| 🞏 A lot | 🞏 Some |
| --- | --- |
| 🞏 A little | 🞏 Not at all |

**7) Are (or were) any of these dears out of proportion to the actual danger involved?**

| 🞏 Yes | 🞏 No |
| --- | --- |

**8) Did you ever try the following for these problems?**

| 🞏 Medication prescribed to  you for at least two weeks | 🞏 Specific anti-anxiety medication prescribed to you for at least one week | 🞏 Unprescribed medication  more than once |
| --- | --- | --- |
| 🞏 Drugs or alcohol more than once | 🞏Psychotherapy or other talking therapy more than once (including internet based CBT) | 🞏Structured wellbeing activity (e.g. mindfulness, meditation, self-help book)_ |
| 🞏Regular physical exercise (e.g. yoga, running, walking) | 🞏 Prefer not to answer | 🞏None of the above |

**Question 8a is only displayed to those who selected ‘psychotherapy or other talking therapy’ in Q8.**

**8a) Are you currently enrolled in an NHS funded talking therapy of psychotherapy (IAPT) for these problems?**

| 🞏 Yes | 🞏 Don’t know |
| --- | --- |
| 🞏 No |  |

**Questions 8b and 8c are only displayed to those who selected ‘prescribed medication’ in Q8.**

**8b) Did you take your medication as advised?**

| 🞏 Yes | 🞏 No |
| --- | --- |
| 🞏 Don’t know | 🞏 Prefer not to answer |

**8c) Did you find the medication helpful?**

| 🞏 Yes | 🞏 No |
| --- | --- |
| 🞏 Don’t know | 🞏 Prefer not to answer |

**Question 9-9b is only displayed to those who selected “psychotherapy or other talking therapy” or “structured wellbeing activity” in Q8**

**9) You previously mentioned that you have tried psychotherapy or another talking therapy, please select all that you attended more than once.**

| 🞏 Counselling | 🞏 Group therapy | 🞏 Cognitive Behavioral Therapy (CBT) |
| --- | --- | --- |
| 🞏 Mindfulness | 🞏 Guided self-help | 🞏 Workshops |
| 🞏 Relationship therapy | 🞏 Family therapy | 🞏 Online therapy |
| 🞏 Never tried psychotherapy or other talking therapies *(skip to next section)* | 🞏 Prefer not to answer | 🞏 Other |

**9a) Did you complete your course of psychotherapy or other talking therapy?**

| 🞏 Yes | 🞏 No |
| --- | --- |
| 🞏 Don’t know | 🞏 Prefer not to answer |

**9b) Did you find psychotherapy or other talking therapy helpful?**

| 🞏 Yes | 🞏 No |
| --- | --- |
| 🞏 Don’t know | 🞏 Prefer not to answer |

#### 1e. Social anxiety disorder (social phobia)

##### DSM-5 Diagnostic criteria

A. Marked fear or anxiety about one or more social situations in which the individual is exposed to possible scrutiny by others. Examples include social interactions (e.g., having a conversation, meeting unfamiliar people), being observed (e.g., eating or drink­ing), and performing in front of others (e.g., giving a speech).

B. The individual fears that he or she will act in a way or show anxiety symptoms that will be negatively evaluated (i.e., will be humiliating or embarrassing: will lead to rejection or offend others).

C. The social situations almost always provoke fear or anxiety.

D. The social situations are avoided or endured with intense fear or anxiety.

E. The fear or anxiety is out of proportion to the actual threat posed by the social situation and to the sociocultural context.

F. The fear, anxiety, or avoidance is persistent, typically lasting for 6 months or more.

G. The fear, anxiety, or avoidance causes clinically significant distress or impairment in social, occupational, or other important areas of functioning.

H. The fear, anxiety, or avoidance is not attributable to the physiological effects of a substance (e.g., a drug of abuse, a medication) or another medical condition.

I. The fear, anxiety, or avoidance is not better explained by the symptoms of another mental disorder, such as panic disorder, body dysmorphic disorder, or autism spectrum disorder.

J. If another medical condition (e.g., Parkinson’s disease, obesity, disfigurement from burns or injury) is present, the fear, anxiety, or avoidance is clearly unrelated or is excessive.

Specify if:

Performance only: If the fear is restricted to speaking or performing in public.

##### Scoring algorithm

**IF**

| Do you have (or have you ever had) a strong fear of, or are (were) you extremely anxious about any of the following situations…   - Being in social situations (e.g. talking with and meeting unfamiliar people)   **Yes** **Is Selected**  **AND/OR**   - Being observed (e.g. eating or drinking while others are watching, talking in front of others)   **Yes** **Is Selected** | **Criterion A**  Marked fear or anxiety about one or more social situations in which the individual is exposed to possible scrutiny by others. Examples include social interactions, being observed, and performing in front of others. |
| --- | --- |
| *Variable name:*  **socp.anx_social_situations** – Yes (1) **AND/OR** **socp.anx_being_observed –** Yes (1) | |

**AND**

| Are/were you worried about what other people will think in these social situations?  **Yes Is Selected** | **Criterion B**  The individual fears that he or she will act in a way or show anxiety symptoms that will be negatively evaluated. |
| --- | --- |
| *Variable name:*  **socp.anx_others_think** – Yes (1) | |

**AND**

| How often do/did these social situations cause fear or anxiety for you?   - **Almost always is selected**   **OR**   - **Always is selected** | **Criterion C**  The social situations almost always provoke fear or anxiety. |
| --- | --- |
| *Variable name:*  **socp.anx_social_situation_frequency** – Almost always (3) **OR** Always (4) | |

**AND**

| Do you (or did you) …   - Avoid social situations?   **Yes is selected**  **AND/OR**   - Endure them with intense anxiety?   **Yes is selected** | **Criterion D**  The social situations are avoided or endured with intense fear or anxiety. |
| --- | --- |
| *Variable name:*  **socp.avoid_social_situations** – Yes (1) *AND/OR* **socp.endure_social_situations** – Yes (1) | |

**AND**

| Is/was your fear or anxiety in social situations out of proportion to the actual threat posed by the situations?  **Yes Is Selected** | **Criterion E**  The fear or anxiety is out of proportion to the actual threat posed by the social situation and to the sociocultural context. |
| --- | --- |
| *Variable name:*  **socp.anx_irrational** – Yes (1) | |

**AND**

| How long was the longest time any of these fears lasted?   - Between 6 and 12 months **Is Selected**   **OR**   - Between 1 and 5 years **Is Selected**   **OR**   - More than 5 years **Is Selected**   **OR**   - All my life / as long as I can remember **Is Selected** | **Criterion F**  The fear, anxiety, or avoidance is persistent, typically lasting for 6 months or more. |
| --- | --- |
| *Variable name:*  **socp.longest_episode** – 6-12 months (2) **OR** 1-5 years (3) **OR** 5+ years (4) **OR** All of my life (5) | |

**AND**

| How much does /did your fear, anxiety or avoidance of social situations interfere with your ability to do your job, have a social life, or interfere with any other important area of your life?   - A lot **is selected**   OR   - Some **is selected** | **Criterion G**  The fear, anxiety, or avoidance causes clinically significant **distress** or **impairment** in social, occupational, or other important areas of functioning. |
| --- | --- |
| *Variable name:*  **socp.interference_with_life** – Some (2) **OR** A lot (3) | |

*(The options for these questions include: ‘a lot,’ ‘some,’ ‘a little’ or ‘none.’ We have drawn the line*

*of clinically significant distress at ‘some’ or ‘a lot.’)*

##### CIDI-SF measure

**The next questions are about things that make some people so afraid that they avoid them or they endure them with intense fear or anxiety.**

**1) Do you have (or have you ever had) a strong fear of, or are (were) you extremely anxious about, any of the following situations?**

|  | **1**  **No** | **2**  **Yes** |
| --- | --- | --- |
| **a. Being in social situations (e.g. talking with and meeting unfamiliar people)** | 🞏 | 🞏 |
| **b. Being observed (e.g. eating or drinking while others are watching, talking in front of others)** | 🞏 | 🞏 |

**If no is selected for both above statements, skip to the next section. If yes is selected for one or more of these statements, continue to question 2.**

**2) Are (or were) you worried about what other people will think in these social situations?**

| 🞏 Yes | 🞏 No |
| --- | --- |

**3) How often do (or did) these social situations cause fear or anxiety for you?**

| 🞏 Always *(continue to Q4)* | 🞏 Almost always *(continue to Q4)* |
| --- | --- |
| 🞏 Some of the time *(skip to section L)* | 🞏 Only one or two times ever *(skip to section L)* |
| 🞏Never*(skip to section L)* |  |

**4) Thinking about the situations that you fear (or feared)**

**Do you (or did you)...?**

|  | **1**  **No** | **2**  **Yes** |
| --- | --- | --- |
| **a. avoid social situations?** | 🞏 | 🞏 |
| **b. endure them with intense anxiety?** | 🞏 | 🞏 |

**5) Is (or was) your fear or anxiety in social situations out of proportion to the actual threat posed by the situations?**

| 🞏 Yes | 🞏 No |
| --- | --- |

**6) How long was the longest time any of these fears lasted?**

| 🞏 Less than 6 months *(skip to Q8)* | 🞏 Between 6 and 12 months |
| --- | --- |
| 🞏 Between 1 and 5 years | 🞏 More than 5 years |
| 🞏All of my life / as long as I can remember *(skip to Q8)* |  |

**7) How many periods of this kind of fear or anxiety have you had in your life lasting 6 or more months?**

| 🞏 One *(skip to Q8)* | 🞏 Two-three |
| --- | --- |
| 🞏 Several | 🞏 All my life / as long as I can remember |
| 🞏 Prefer not to answer *(skip to Q8)* |  |

**7a) Please estimate the number of times you have had periods of this kind of worry in your life lasting 6 or more months:**

🞏🞏

**8) How old were you when these fears first started?**

🞏🞏

**Question 9 is not displayed to those who selected ‘one’ or ‘prefer not to answer’ in Q7**

**9) How old were you when you most recently experienced these fears?**

🞏🞏

**10) How much have any of these fears ever interfered with your life or activities?**

| 🞏 A lot | 🞏 Some |
| --- | --- |
| 🞏 A little | 🞏 Not at all |

**11) How much does (or did) your fear, anxiety or avoidance of social situations interfere with your ability to do your job, have a social life, or interfere with any other important area of your life?**

| 🞏 A lot | 🞏 Some |
| --- | --- |
| 🞏 A little | 🞏 Not at all |

**12) Did you ever try the following for these problems?**

| 🞏 Medication prescribed to  you for at least two weeks | 🞏 Specific anti-anxiety medication prescribed to you for at least one week | 🞏 Unprescribed medication  more than once |
| --- | --- | --- |
| 🞏 Drugs or alcohol more than once | 🞏Psychotherapy or other talking therapy more than once (including internet based CBT) | 🞏Structured wellbeing activity (e.g. mindfulness, meditation, self-help book)_ |
| 🞏Regular physical exercise (e.g. yoga, running, walking) | 🞏 Prefer not to answer | 🞏None of the above |

**Question 12a is only displayed to those who selected ‘psychotherapy or other talking therapy’ in Q12.**

**12a) Are you currently enrolled in an NHS funded talking therapy of psychotherapy (IAPT) for these problems?**

| 🞏 Yes | 🞏 Don’t know |
| --- | --- |
| 🞏 No |  |

**Questions 12b and 12c are only displayed to those who selected “prescribed medication” in Q12.**

**12b) Did you take your medication as advised?**

| 🞏 Yes | 🞏 No |
| --- | --- |
| 🞏 Don’t know | 🞏 Prefer not to answer |

**12b) Did you find the medication helpful?**

| 🞏 Yes | 🞏 No |
| --- | --- |
| 🞏 Don’t know | 🞏 Prefer not to answer |

**Question 13 is only displayed to those who selected “psychotherapy or other talking therapy” or ‘structured wellbeing activity’ in question 12**

**13) You previously mentioned that you have tried psychotherapy, another talking therapy, or a structured wellbeing activity, please select all that you attended more than once.**

| 🞏 Counselling | 🞏 Group therapy | 🞏 Cognitive Behavioral Therapy (CBT) |
| --- | --- | --- |
| 🞏 Mindfulness | 🞏 Guided self-help | 🞏 Workshops |
| 🞏 Relationship therapy | 🞏 Family therapy | 🞏 Online therapy |
| 🞏 Never tried psychotherapy or other talking therapies | 🞏 Prefer not to answer | 🞏 Other |

**Q13a-b is only displayed to those who selected “psychotherapy or other talking therapy” in Q12**

**13a) Did you complete your course of psychotherapy or other talking therapy?**

| 🞏 Yes | 🞏 No |
| --- | --- |
| 🞏 Don’t know | 🞏 Prefer not to answer |

**13b) Did you find psychotherapy or other talking therapy helpful?**

| 🞏 Yes | 🞏 No |
| --- | --- |
| 🞏 Don’t know | 🞏 Prefer not to answer |

#### 1f. Panic Disorder

##### DSM-5 Diagnostic criteria

A. Recurrent unexpected panic attacks. A panic attack is an abrupt surge of intense fear or intense discomfort that reaches a peak within minutes, and during which time four (or more) of the following symptoms occur; Note: The abrupt surge can occur from a calm state or an anxious state.

1. Palpitations, pounding heart, or accelerated heart rate.
2. Sweating.
3. Trembling or shaking.
4. Sensations of shortness of breath or smothering.
5. Feelings of choking.
6. Chest pain or discomfort.
7. Nausea or abdominal distress.
8. Feeling dizzy, unsteady, light-headed, or faint.
9. Chills or heat sensations.
10. Paresthesias (numbness or tingling sensations).
11. Derealization (feelings of unreality) or depersonalization (being detached from one­self).
12. Fear of losing control or “going crazy.”
13. Fear of dying.

Note: Culture-specific symptoms (e.g., tinnitus, neck soreness, headache, uncontrol­lable screaming or crying) may be seen. Such symptoms should not count as one of the four required symptoms.

B. At least one of the attacks has been followed by 1 month (or more) of one or both of the following:

1. Persistent concern or worry about additional panic attacks or their consequences (e.g., losing control, having a heart attack, “going crazy”).
2. A significant maladaptive change in behavior related to the attacks (e.g., behaviors designed to avoid having panic attacks, such as avoidance of exercise or unfamiliar situations).

C. The disturbance is not attributable to the physiological effects of a substance (e.g., a drug of abuse, a medication) or another medical condition (e.g., hyperthyroidism, car­diopulmonary disorders).

D. The disturbance is not better explained by another mental disorder (e.g., the panic at­tacks do not occur only in response to feared social situations, as in social anxiety dis­order: in response to circumscribed phobic objects or situations, as in specific phobia: in response to obsessions, as in obsessive-compulsive disorder: in response to re­minders of traumatic events, as in posttraumatic stress disorder: or in response to sep­aration from attachment figures, as in separation anxiety disorder)

##### Scoring algorithm

**IF**

| Have you ever had a sudden, unexpected surge of intense fear or intense discomfort (panic attack) during which you experienced any of the following symptoms…   - Your heart was pounding or racing - You were sweating - You were trembling or shaking - You felt short of breath, or like you were being smothered - You felt like you were choking - You had pain or discomfort in your chest - You were nauseous or felt sick in the stomach - You felt dizzy, unsteady, light-headed or faint - You felt hot or cold - You felt numbness or tingling sensations - It felt like things weren't real, or you felt detached from yourself - You were afraid you were going to lose control or "go crazy" - You were afraid you were going to die - No, I have never had this happen to me   **At least four symptoms are selected** | **Criterion A**  Recurrent unexpected panic attacks. A panic attack is an abrupt surge of intense fear or intense discomfort that reaches a peak within minutes, and during which time four (or more) of the following symptoms occur: |
| --- | --- |
| *Variable name:*  **At least four of the following:**  **pad.heart_pounding –** Yes (1) *AND/OR* **pad.sweating –** Yes (1) *AND/OR* **pad.trembling –** Yes (1)  *AND/OR* **pad.short_of_breath –** Yes (1) *AND/OR* **pad.choking –** Yes (1) *AND/OR*  **pad.chest_pain –** Yes (1) *AND/OR* **pad.nauseous –** Yes (1) *AND/OR* **pad.dizzy –** Yes (1)  *AND/OR* **pad.hot_cold –** Yes (1) *AND/OR* **pad.numbness –** Yes (1) *AND/OR*  **pad.detached –** Yes (1) *AND/OR* **pad.lose_control –** Yes (1) *AND/OR* **pad.going_to_die –** Yes (1) | |

**AND**

| After any of your attacks of fear or panic, did you ever…   - Feel anxious, worried or nervous about having more panic attacks? **Yes is selected**   *AND/OR*   - Feel worried about losing control, having a heart attack, going crazy, or other bad things happening because of panic attacks? **Yes is selected**   *AND/OR*   - Avoid situations in which panic attacks might occur? **Yes is selected** | **Criterion B**  At least one of the attacks has been followed by 1 month (or more) of one or both of the following:   1) Persistent concern or worry about additional panic attacks or their consequences (e.g., losing control, having a heart attack, “going crazy”). 2) A significant maladaptive change in behavior related to the attacks (e.g., behaviors designed to avoid having panic attacks, such as avoidance of exercise or unfamiliar situations). |
| --- | --- |
| *Variable name:*  **pad.anx_future_panic_attacks** – Yes (1) *AND/OR* **pad.worried_future_panic_attacks** – Yes (1) *AND/OR* **pad.avoid_situation_panic_attacks** – Yes (1) | |

**AND**

| How long did you continue to worry about panic attacks or their consequences, or avoid situations in which panic attacks might occur?   - Between 1 and 6 months **Is Selected**   **OR**   - Between 6 and 12 months **Is Selected**   **OR**   - More than 12 months **Is Selected**   **OR**   - Between 1 and 5 years **Is Selected**   **OR**   - More than 5 years **Is Selected**   **OR**   - All of my life/as long as I can remember **Is Selected** | **Criterion B**  At least one of the attacks has been followed by **1 month (or more)** of one or both of the following:   1) Persistent concern or worry about additional panic attacks or their consequences (e.g., losing control, having a heart attack, “going crazy”). 2) A significant maladaptive change in behavior related to the attacks (e.g., behaviors designed to avoid having panic attacks, such as avoidance of exercise or unfamiliar situations). |
| --- | --- |
| *Variable name:*  **pad.duration** – 1-6 months (2) **OR** 6-12 months (3) **OR** 1-5 years (4) **OR** 5+ years (5) **OR**  All of my life – (6) **OR** 12+ months – (7)* | |

**12+ months response option was replaced partway through data collection with options 4-6 to collect more detailed information about symptom duration*

**AND**

| Were these attacks or sudden periods of physical discomfort ever the result of a medical condition (e.g. a heart attack) or from using medication, drugs or alcohol?   - **‘No, never’ is selected**   **OR**   - **Yes, some of them is selected** | **Criterion C**  The disturbance is not attributable to the physiological effects of a substance (e.g., a drug of abuse, a medication) or another medical condition (e.g., hyperthyroidism, cardiopulmonary disorders). |
| --- | --- |
| *Variable name:*  **pad.physical_cause** – No, never (0) **OR** Yes, some of them (1) | |

**AND**

| We already asked about specific situations that cause strong fears (heights, elevators, snakes etc). When you have sudden anxiety attacks, do they **usually** occur in specific situations that cause you strong fear?  **No is selected**  *AND/OR*  Did you ever have an attack when you were **not** in a situation that usually causes you to have strong fears?  **Yes is selected** | **Criterion D**  The disturbance is not better explained by another mental disorder (e.g., the panic attacks do not occur only in response to feared social situations, as in social anxiety disorder; in response to circumscribed phobic objects or situations, as in specific phobia; in response to obsessions, as in obsessive-compulsive disorder; in response to reminders of traumatic events, as in posttraumatic stress disorder; or in response to separation from attachment figures, as in separation anxiety disorder). |
| --- | --- |
| *Variable name:*  **pad.heights** – No (0) *AND/OR* **pad.random** – Yes (1) | |

##### CIDI-SF measure

**The next questions relate to any experiences you may have had with panic attacks or feelings of intense panic**

**Have you ever had a sudden, unexpected surge of intense fear or intense discomfort (panic attack) during which you experienced some of the following symptoms?  

(Please select all symptoms that occurred at the same time)**

| 🞏 Your heart was pounding or racing | 🞏 You were sweating |
| --- | --- |
| 🞏You were trembling or shaking | 🞏 You felt short of breath, or like you were being smothered |
| 🞏You felt like you were choking | 🞏You had pain or discomfort in your chest |
| 🞏You were nauseous or felt sick in the stomach | 🞏You felt dizzy, unsteady, light-headed or faint |
| 🞏You felt hot or cold | 🞏You felt numbness or tingling sensations |
| 🞏It felt like things weren't real, or you felt detached from yourself | 🞏You were afraid you were going to lose control or "go crazy" |
| 🞏You were afraid you were going to die | 🞏No, I have never had this happen to me |

**If less than three of the above statements are selected please skip to the next section. If three or more of the above statements are selected, please continue to question 2.**

**2) How many such attacks of fear or panic would you say that you have had over the course of your lifetime?**

🞏🞏

**3)** **After any of your attacks of fear or panic, did you ever ...**

|  | **1**  **No** | **2**  **Yes** |
| --- | --- | --- |
| **a. Feel anxious, worried or nervous about having more panic attacks?** | 🞏 | 🞏 |
| **b. Feel worried about losing control, having a heart attack, going crazy, or other bad things happening because of panic attacks?** | 🞏 | 🞏 |
| **c. Avoid situations in which panic attacks might occur?** | 🞏 | 🞏 |

**4) How long did you continue to worry about panic attacks or their consequences, or avoid situations in which panic attacks might occur?**

| 🞏 Less than 1 month *(skip to 6)* | 🞏 Over 1 month but less than 6 months |
| --- | --- |
| 🞏 Over 6 months but less than 12 months | 🞏 Over 1 year but less than 5 years |
| 🞏 More than 5 years | 🞏All of my life / as long as I can remember |

**5) How many periods of this kind of worry have you had in your life lasting 1 or more months?**

| 🞏 One *(skip to Q6)* | 🞏 Two-three |
| --- | --- |
| 🞏 Several | 🞏 All my life/ as long as I can remember |
| 🞏Prefer not to answer *(skip to Q6)* |  |

**5a) Please estimate the number of times you have had periods of this kind of worry in your life lasting 1 or more months:**

🞏🞏

**6) Were these attacks or sudden periods of physical discomfort ever the result of a medical condition (e.g. a heart attack) or from using medication, drugs or alcohol?**

| 🞏 No, never | 🞏 Yes, some of them |
| --- | --- |
| 🞏 Yes, all of them |  |

**7) We already asked about specific situations that cause strong fears (heights, elevators, snakes etc). When you have sudden anxiety attacks, do they usually occur in specific situations that cause you strong fear?**

| 🞏 No | 🞏 Yes |
| --- | --- |

**8) Did you ever have an attack when you were not in a situation that usually causes you to have strong fears?**

| 🞏 No | 🞏 Yes |
| --- | --- |

**9) How old were you the first time you had one of these sudden attacks of feeling frightened, anxious or panicky?**

🞏🞏

**10) How old were you the last time you had one of these sudden attacks of feeling frightened, anxious or panicky?**

🞏🞏

**11) Did you ever try the following for these problems?**

| 🞏 Medication prescribed to  you for at least two weeks | 🞏 Specific anti-anxiety medication prescribed to you for at least one week | 🞏 Unprescribed medication  more than once |
| --- | --- | --- |
| 🞏 Drugs or alcohol more than once | 🞏Psychotherapy or other talking therapy more than once (including internet based CBT) | 🞏Structured wellbeing activity (e.g. mindfulness, meditation, self-help book) |
| 🞏Regular physical exercise (e.g. yoga, running, walking) | 🞏 Prefer not to answer | 🞏None of the above |

**Question 11a is only displayed to those who selected ‘psychotherapy or other talking therapy’ in Q11.**

**11a) Are you currently enrolled in an NHS funded talking therapy of psychotherapy (IAPT) for these problems?**

| 🞏 Yes | 🞏 Don’t know |
| --- | --- |
| 🞏 No |  |

**Questions 11b and 11c are only displayed to those who selected “prescribed medication” in question 10.**

**11b) Did you take your medication as advised?**

| 🞏 Yes | 🞏 No |
| --- | --- |
| 🞏 Don’t know | 🞏 Prefer not to answer |

**11c) Did you find the medication useful?**

| 🞏 Yes | 🞏 No |
| --- | --- |
| 🞏 Don’t know | 🞏 Prefer not to answer |

**Q12 is only displayed to those who selected ‘psychotherapy or other talking therapy’ or ‘structured wellbeing activity’ in Q11**

**12) You previously mentioned that you have tried psychotherapy, another talking therapy, or structured wellbeing activity. Please select all that you attended more than once.**

| 🞏 Counselling | 🞏 Group therapy | 🞏 Cognitive Behavioral Therapy (CBT) |
| --- | --- | --- |
| 🞏 Mindfulness | 🞏 Guided self-help | 🞏 Workshops |
| 🞏 Relationship therapy | 🞏 Family therapy | 🞏 Online therapy |
| 🞏 Never tried psychotherapy or other talking therapies | 🞏 Prefer not to answer | 🞏 Other |

**Q12a-b is only displayed to those who selected ‘psychotherapy or other talking therapy’ in Q11**

**12a) Did you complete your course of psychotherapy or other talking therapy?**

| 🞏 Yes | 🞏 No |
| --- | --- |
| 🞏 Don’t know | 🞏 Prefer not to answer |

**12b) Did you find psychotherapy or other talking therapy useful?**

| 🞏 Yes | 🞏 No |
| --- | --- |
| 🞏 Don’t know | 🞏 Prefer not to answer |

#### 1g. Agoraphobia

##### DSM-5 Diagnostic criteria

A. Marked fear or anxiety about two (or more) of the following five situations:

Using public transportation (e.g., automobiles, buses, trains, ships, planes).

Being in open spaces (e.g., parking lots, marketplaces, bridges).

Being in enclosed places (e.g., shops, theaters, cinemas).

Standing in line or being in a crowd.

Being outside of the home alone.

B. The individual fears or avoids these situations because of thoughts that escape might be difficult or help might not be available in the event of developing panic-like symp­toms or other incapacitating or embarrassing symptoms (e.g., fear of falling in the el­derly; fear of incontinence).

C. The agoraphobic situations almost always provoke fear or anxiety.

D. The agoraphobic situations are actively avoided, require the presence of a companion, or are endured with intense fear or anxiety.

E. The fear or anxiety is out of proportion to the actual danger posed by the agoraphobic situations and to the sociocultural context.

F. The fear, anxiety, or avoidance is persistent, typically lasting for 6 months or more.

G. The fear, anxiety, or avoidance causes clinically significant distress or impairment in social, occupational, or other important areas of functioning.

H. If another medical condition (e.g., inflammatory bowel disease, Parkinson’s disease) is present, the fear, anxiety, or avoidance is clearly excessive.

I. The fear, anxiety, or avoidance is not better explained by the symptoms of another men­tal disorder—for example, the symptoms are not confined to specific phobia, situational type; do not involve only social situations (as in social anxiety disorder): and are not re­lated exclusively to obsessions (as in obsessive-compulsive disorder), perceived defects or flaws in physical appearance (as in body dysmorphic disorder), reminders of traumatic events (as in posttraumatic stress disorder), or fear of separation (as in separation anx­iety disorder).

Note: Agoraphobia is diagnosed irrespective of the presence of panic disorder. If an indi­vidual’s presentation meets criteria for panic disorder and agoraphobia, both diagnoses should be assigned

##### Scoring algorithm

**IF**

| Do you have (or have you ever had) a strong fear of, or are (were) you extremely anxious about, any of the following situations…  **At least two situations are selected**   - Using public transportation (e.g. cars, buses, trains, ships, planes) - Being in open spaces (e.g. parking lots, marketplaces, bridges) - Being in enclosed spaces (e.g. shops, theatres, cinemas) - Standing in line or being in a crowd - Being outside of the home alone | **Criterion A**  Marked fear or anxiety about two (or more) of the following five situations: |
| --- | --- |
| *Variable name:*  **At least two of the following:**  **agp.public_transport_phobia –** Yes (1) *AND/OR* **agp.open_spaces_phobia –** Yes (1) *AND/OR*  **agp.enclosed_spaces_phobia –** Yes (1) *AND/OR* **agp.queue_or_crowd_phobia –** Yes (1) *AND/OR*  **agp.outside_home_alone_phobia –** Yes (1) | |

**AND**

| - In one or more of these situations, are/were you ever afraid that you might faint, lose control, or embarrass yourself in other ways?   **Yes is selected**  *AND/OR*   - Are/were you afraid that escape might be difficult if that happened?   **Yes is selected**  *AND/OR*   - Are/were you afraid that help might not be available if you needed it?   **Yes is selected** | **Criterion B**  The individual fears or avoids these situations because of thoughts that escape might be difficult or help might not be available in the event of developing panic-like symptoms or other incapacitating or embarrassing symptoms (e.g., fear of falling in the elderly; fear of incontinence). |
| --- | --- |
| *Variable name:*  **agp.afraid_faint** – Yes (1) *AND/OR* **agp.afraid_escape_difficult** – Yes (1) *AND/OR*  **agp.afraid_help_not_available** – Yes (1) | |

**AND**

| How often do/did these situations cause fear or anxiety for you?   - **Almost always is selected**   **OR**   - **Always is selected** | **Criterion C**  The agoraphobic situations almost always provoke fear or anxiety. |
| --- | --- |
| *Variable name:*  **agp.phobia_frequency** – Almost always (3) **OR** Always (4) | |

**AND**

| Do you (or did you) …   - Avoid these situations? **Yes is selected**   **AND/OR**   - Endure them with intense anxiety? **Yes is selected**   **AND/OR**   - Require the presence of a companion? **Yes** **is selected** | **Criterion D**  The agoraphobic situations are actively avoided, require the presence of a companion, or are endured with intense fear or anxiety. |
| --- | --- |
| *Variable name:*  **agp.avoid_phobia** – Yes (1) *AND/OR* **agp.endure_phobia_with_anxiety** – Yes (1) *AND/OR* **agp.require_companion** – Yes (1) | |

**AND**

| Are (or were) any of these fears out of proportion to the actual danger involved?  **Yes is selected** | **Criterion E**  The fear or anxiety is out of proportion to the actual danger posed by the agoraphobic situations and to the sociocultural context. |
| --- | --- |
| *Variable name:*  **agp.phobia_out_of_proportion** – Yes (1) | |

**AND**

| How long was the longest time any of these fears lasted?   - Between 6 and 12 months **Is Selected**   **OR**   - Between 1 and 5 years **Is Selected**   **OR**   - All of my life / As long as I can remember **Is Selected** | **Criterion F**  The fear, anxiety, or avoidance is persistent, typically lasting for 6 months or more. |
| --- | --- |
| *Variable name:*  **agp.phobia_lasted** – 6-12 months (2) **OR** 1-5 years (3) **OR** 5+ years (4) **OR** All of my life (5) | |

**AND**

| How much have any of these fears ever interfered with your life or activities?   - **A lot Is Selected**   **OR**   - **Some Is Selected** | **Criterion G**  The fear, anxiety, or avoidance causes clinically significant distress or impairment in social, occupational, or other important areas of functioning. |
| --- | --- |
| *Variable name:*  **agp.phobia_interfered** – Some (2) **OR** A lot (3) | |

*(The options for these questions include: ‘a lot,’ ‘some,’ ‘a little’ or ‘none.’ We have drawn the line*

*of clinically significant distress at ‘some’ or ‘a lot.’)*

##### CIDI-SF measure

**The next questions contain a list of situations which some people actively avoid, need a companion with them for, or endure with intense fear or anxiety.**

**1) Do you have (or have you ever had) a strong fear of, or are (were) you extremely anxious about, any of the following situations?**

|  | **1**  **No** | **2**  **Yes** |
| --- | --- | --- |
| **a. Using public transportation (e.g. cars, buses, trains, ships, planes)** | 🞏 | 🞏 |
| **b. Being in open spaces (e.g. parking lots, marketplaces, bridges)** | 🞏 | 🞏 |
| **c. Being in enclosed spaces (e.g. shops, theatres, cinemas)** | 🞏 | 🞏 |
| **d. Standing in line or being in a crowd** | 🞏 | 🞏 |
| **e. Being outside of the home alone** | 🞏 | 🞏 |

**If yes is selected for two or more of these statements, please continue to question 2**

**If no is selected for four or all of the above statements please skip to the next section.**

**2) How often do (or did) these social situations cause fear or anxiety for you?**

| 🞏 Always *(continue to Q3)* | 🞏 Almost always *(continue to Q3)* |
| --- | --- |
| 🞏 Some of the time *(continue to Q3)* | 🞏 Only one or two times ever *(Continue to Q3)* |
| 🞏Never*(skip to next section)* |  |

**2.1) Do you (or did you)...?**

|  | **1**  **No** | **2**  **Yes** |
| --- | --- | --- |
| **a. Avoid social situations?** | 🞏 | 🞏 |
| **b. Endure them with intense anxiety?** | 🞏 | 🞏 |
| **c. Require the presence of a companion?** | 🞏 | 🞏 |

**3) Thinking about the situations that you fear (or feared)**

|  | **1**  **No** | **2**  **Yes** |
| --- | --- | --- |
| **a. In one or more of these situations, are (were) you ever afraid that you might faint, lose control, or embarrass yourself in other ways?** | 🞏 | 🞏 |
| **b. Are (were) you afraid that escape might be difficult if that happened?** | 🞏 | 🞏 |
| **c. Are (were) you afraid that help might not be available if you needed it?** | 🞏 | 🞏 |

**4) How old were you when these fears first started?**

🞏🞏

**5) How old were you when you most recently experienced any of these fears?**

🞏🞏

**6) How long was the longest time any of these fears lasted?**

| 🞏 Less than 6 months (skip to 6) | 🞏 Between 6 and 12 months |
| --- | --- |
| 🞏 Between 1 and 5 years | 🞏 More than 5 years |
| 🞏All of my life / as long as I can remember (skip to 6) |  |

**7) How many periods of this kind of fear or anxiety have you had in your life lasting 6 or more months?**

| 🞏 One (skip to 6) | 🞏 Two-three (continue to 5a) |
| --- | --- |
| 🞏 Several (continue to 5a) | 🞏 All my life/ as long as I can remember (skip to 6) |
| 🞏Prefer not to answer (skip to Q6) |  |

**7a) Please estimate the number of times you have had periods of this kind of fear or anxiety in your life lasting 6 or more months:**

🞏🞏

**8) How much have any of these fears ever interfered with your life or activities?**

| 🞏 A lot | 🞏 Some |
| --- | --- |
| 🞏 A little | 🞏 Not at all |

**9) Are (or were) any of these fears out of proportion to the actual danger involved?**

| 🞏 Yes | 🞏 No |
| --- | --- |

**10) Did you ever try the following for these problems?**

| 🞏 Medication prescribed to  you for at least two weeks | 🞏 Specific anti-anxiety medication prescribed to you for at least one week | 🞏 Unprescribed medication  more than once |
| --- | --- | --- |
| 🞏 Drugs or alcohol more than once | 🞏Psychotherapy or other talking therapy more than once (including internet based CBT) | 🞏Structured wellbeing activity (e.g. mindfulness, meditation, self-help book) |
| 🞏Regular physical exercise (e.g. yoga, running, walking) | 🞏 Prefer not to answer | 🞏None of the above |

**Question 10a is only displayed to those who selected ‘psychotherapy or other talking therapy’ in Q10.**

**10a) Are you currently enrolled in an NHS funded talking therapy of psychotherapy (IAPT) for these problems?**

| 🞏 Yes | 🞏 Don’t know |
| --- | --- |
| 🞏 No |  |

**Q10b and 10c are only displayed to those who selected “prescribed medication” in question Q10.**

**10b) Did you take your medication as advised?**

| 🞏 Yes | 🞏 No |
| --- | --- |
| 🞏 Don’t know | 🞏 Prefer not to answer |

**10c) Did you find the medication helpful?**

| 🞏 Yes | 🞏 No |
| --- | --- |
| 🞏 Don’t know | 🞏 Prefer not to answer |

**Question 11 is only displayed to those who selected “psychotherapy or other talking therapy” or ‘structured wellbeing activity’ in Q10**

**11) You previously mentioned that you have tried psychotherapy, another talking therapy, or a structured wellbeing activity. please select all that you attended more than once.**

| 🞏 Counselling | 🞏 Group therapy | 🞏 Cognitive Behavioral Therapy (CBT) |
| --- | --- | --- |
| 🞏 Mindfulness | 🞏 Guided self-help | 🞏 Workshops |
| 🞏 Relationship therapy | 🞏 Family therapy | 🞏 Online therapy |
| 🞏 Never tried psychotherapy or other talking therapies | 🞏 Prefer not to answer | 🞏 Other |

**Q11a-b are only displayed to those who selected “psychotherapy or other talking therapy” in Q10**

**11a) Did you complete your course of psychotherapy or other talking therapy?**

| 🞏 Yes | 🞏 No |
| --- | --- |
| 🞏 Don’t know | 🞏 Prefer not to answer |

**11b) Did you find psychotherapy or other talking therapy helpful?**

| 🞏 Yes | 🞏 No |
| --- | --- |
| 🞏 Don’t know | 🞏 Prefer not to answer |

### Appendix 2: Single-item diagnoses

Single-time diagnoses were identified by responses on a single self-report question. The exact phrasing of this question and how responses were mapped to disorders is detailed below.

“Have you **ever been diagnosed** with one or more of the following mental health problems by a **professional**, even if you don't have it currently?”

| **Response option** | **Single-item diagnosis** |
| --- | --- |
| Depression | Major depressive disorder |
| Anxiety, nerves or generalised anxiety disorder | Generalised anxiety disorder |
| Specific phobia *(e.g. phobia of flying)* | Specific phobia |
| Social anxiety or social phobia | Social anxiety disorder |
| Panic disorder | Panic disorder |
| Panic attacks | Panic attacks |
| Agoraphobia | Agoraphobia |

### Appendix 3: Missing data

#### Appendix 3a. Number of participants with missing data for each variable

| **Variable** | **NA Count** | **Percent** |
| --- | --- | --- |
| Ethnicity | 3996 | 6.84 |
| Highest education | 3186 | 5.46 |
| Employment | 11681 | 20.00 |
| Single-item MDD | 1645 | 2.82 |
| Single-item GAD | 1645 | 2.82 |
| Single-item specific phobia | 1645 | 2.82 |
| Single-item social anxiety disorder | 1645 | 2.82 |
| Single-item panic attacks | 1645 | 2.82 |
| Single-item panic disorder | 14999 | 25.68 |
| Single-item agoraphobia | 1645 | 2.82 |
| Single-item any anxiety | 4241 | 7.26 |
| Algorithm-based MDD | 5326 | 9.12 |
| Algorithm-based GAD | 13659 | 23.39 |
| Algorithm-based specific phobia | 10278 | 17.60 |
| Algorithm-based social anxiety disorder | 10344 | 17.71 |
| Algorithm-based panic disorder | 11190 | 19.16 |
| Algorithm-based agoraphobia | 11348 | 19.43 |
| Algorithm-based any anxiety | 13709 | 23.47 |

The table displays the number (NA Count) and percentage of participants with missing data on each variable from the full sample (N = 58,400).

The counts are not exclusive as participants may have missing data on multiple variables above.

Abbreviations: MDD, major depressive disorder; GAD, generalised anxiety disorder

#### Appendix 3b. Frequencies of the number of missing algorithm-based diagnoses.

| **Number of missing**  **AB diagnoses** | **Frequency** | **Percent** |
| --- | --- | --- |
| 0 | 36601 | 62.67 |
| 1 | 10425 | 17.85 |
| 2 | 1017 | 1.741 |
| 3 | 185 | 0.32 |
| 4 | 4516 | 7.73 |
| 5 | 2869 | 4.91 |
| 6 | 2787 | 4.77 |
| Total | 58400 | 100.00 |

The table displays the frequencies of participants with 0-6 missing algorithm-based diagnoses from the full sample (N = 58,400).

Abbreviations: AB, Algorithm-based

#### Appendix 3c. Associations between demographics and missing data for algorithm-based and single-item diagnoses.

##### Table 3c-1. Associations between demographic variables and missing data on algorithm-based diagnoses.

| **Characteristic** | **Beta** | **95% CI***^1^* | **p-value** |
| --- | --- | --- | --- |
| Age | 0.01 | 0.005, 0.01 | <0.001*** |
| Sex | -0.14 | -0.17, -0.11 | <0.001*** |
| Ethnicity |  |  |  |
| White | — | — |  |
| Mixed | 0.12 | 0.02, 0.22 | 0.021* |
| Asian or Asian British | 0.43 | 0.30, 0.55 | <0.001*** |
| Black or Black British | 0.20 | -0.01, 0.42 | 0.062 |
| Arab | 0.34 | -0.22, 0.89 | 0.2 |
| Other | -0.16 | -0.34, 0.02 | 0.078 |
| Education |  |  |  |
| GCSE/CSE | — | — |  |
| NVQ | -0.01 | -0.08, 0.05 | 0.7 |
| A-levels | -0.14 | -0.20, -0.09 | <0.001*** |
| University | -0.20 | -0.25, -0.16 | <0.001*** |
| ^1^ CI = Confidence Interval | | | |

*Model fit statistics:*

Residual standard error: 1.713 on 51849 degrees of freedom

(6540 observations deleted due to missingness)

Multiple R-squared: 0.007976,	Adjusted R-squared: 0.007785

F-statistic: 41.69 on 10 and 51849 DF; p-value: < 2.2e-16

The table displays results from a linear regression model. The outcome variable was the sum of missing algorithm-based diagnoses (range 0-6). Frequencies of participants with 0-6 missing algorithm-based diagnoses is displayed in Appendix 4b. Although p-value demonstrated significant differences found for age due to the large sample size, the effect size was too small (0.01) to reflect a relevant difference.

##### Table 3c-2. Associations between demographic variables and missing data on single-item diagnoses.

| **Characteristic** | **Beta** | **95% CI***^1^* | **p-value** |
| --- | --- | --- | --- |
| Age | -3.2e-04 | -0.0004, -0.0002 | <0.001*** |
| Sex | 7.8e-05 | -0.003, 0.003 | >0.9 |
| Ethnicity |  |  |  |
| White | — | — |  |
| Mixed | 0.01 | -0.003, 0.01 | 0.2 |
| Asian or Asian British | 0.01 | -0.005, 0.02 | 0.3 |
| Black or Black British | 3.2e-03 | -0.01, 0.02 | 0.7 |
| Arab | 4.9e-03 | -0.04, 0.05 | 0.8 |
| Other | 0.02 | 0.0005, 0.03 | 0.042* |
| Highest education |  |  |  |
| GCSE/CSE | — | — |  |
| NVQ | 5.0e-04 | -0.005, 0.01 | 0.8 |
| A-levels | -0.01 | -0.01, -0.005 | <0.001*** |
| University | -0.01 | -0.01, -0.01 | <0.001*** |
| ^1^ CI = Confidence Interval | | | |

*Model fit statistics:*

Null deviance: 1024.9 on 51859 degrees of freedom

Residual deviance: 1022.8 on 51849 degrees of freedom

(6540 observations deleted due to missingness)

AIC: -56406

The table displays results from a logistic regression model. Due to the method of data collection for single-item diagnoses, the outcome was a binary variable (0-1) for participants missing none (N = 56,755) or all (N = 1,645) single-item diagnoses. Panic disorder was included partway through data collection so participants with missing data on panic disorder only (N = 13,354) were categorised as having no missing data. Although p-values demonstrated significant differences found for some characteristics due to the large sample size, effect sizes were too small (<0.03) to reflect meaningful differences.

*p < 0.05, **p < 0.01, ***p < 0.001

### Appendix 4: Agreement and disagreement on algorithm-based vs single-item diagnoses by sex.

#### Appendix 4a. Figure displaying the agreement and disagreement between the measures by sex.

|  | Agreement: Algorithm-based = Yes, Single-item = Yes | Agreement: Algorithm-based = No, Single-item = No | Disagreement: Algorithm-based = Yes, Single-item = No | Disagreement: Algorithm-based = No, Single-item = Yes |
| --- | --- | --- | --- | --- |
| Agora- phobia Male | 2.71674% | 78.4645% | 17.4181% | 1.40068% |
| Agora- phobia Female | 2.89349% | 77.1021% | 18.5669% | 1.43755% |
| Panic disorder Male | 2.9487% | 73.0757% | 18.6572% | 5.31843% |
| Panic disorder Female | 3.13691% | 71.801% | 19.4897% | 5.57243% |
| Panic attacks† Male | 13.4689% | 51.8873% | 8.13698% | 26.5069% |
| Panic attacks† Female | 14.4143% | 49.2572% | 8.21229% | 28.1162% |
| Social anxiety disorder Male | 16.1608% | 54.1405% | 21.1532% | 8.5455% |
| Social anxiety disorder Female | 16.2022% | 53.6227% | 21.94% | 8.23508% |
| Specific phobia Male | 2.6422% | 79.6156% | 15.8472% | 1.89503% |
| Specific phobia Female | 3.01235% | 77.7322% | 17.1662% | 2.08927% |
| GAD Male | 43.5084% | 14.6404% | 6.64165% | 35.2095% |
| GAD Female | 44.3846% | 13.7117% | 6.29218% | 35.6115% |
| Any anxiety Male | 70.1049% | 6.5766% | 7.03128% | 16.2872% |
| Any anxiety Female | 71.0998% | 6.13255% | 6.97356% | 15.7941% |
| MDD Male | 80.8147% | 3.79152% | 7.84777% | 7.54604% |
| MDD Female | 81.4097% | 3.5349% | 7.75755% | 7.29787% |

Each bar displays the proportions (%) of the full sample by sex (Male: N = 15,786; Female: N = 42,614) with agreement or disagreement between the two measures for each disorder. Agreements are represented in blue (dark blue = agreement on diagnosis, light blue = agreement on no diagnosis) while disagreements are in yellow (dark yellow = algorithm-based but no single-item diagnosis, light yellow = single-item but no algorithm-based diagnosis). †The panic attacks column displays the agreement between algorithm-based panic disorder and single-item panic attacks.

Abbreviations: MDD, major depressive disorder; GAD, generalised anxiety disorder.

#### Appendix 4b: Chi-squared results for differences in agreement and disagreement by sex.

##### Table 4b-1. Chi-squared results comparing differences by sex for categorical agreement and disagreement results for each disorder.

| **disorder** | **statistic** | **df** | **p-value** |
| --- | --- | --- | --- |
| MDD | 2903.1735 | 3 | p < 2.2e-16*** |
| Any anxiety | 2334.5703 | 3 | p < 2.2e-16*** |
| GAD | 2360.7732 | 3 | p < 2.2e-16*** |
| Specific phobia | 925.5512 | 3 | p < 2.2e-16*** |
| Social anxiety disorder | 539.6759 | 3 | p < 2.2e-16*** |
| Panic attacks | 1632.9041 | 3 | p < 2.2e-16*** |
| Panic disorder | 1143.3155 | 3 | p < 2.2e-16*** |
| Agoraphobia | 546.9965 | 3 | p < 2.2e-16*** |

Table 4b-1 displays the result of chi-squared analyses comparing the agreement and disagreement between algorithm-based (AB) and single-item (SI) diagnoses for males (N = 15,786) and females (N = 42,614) for each disorder. The outcome included all 4 levels of agreement/disagreement: agreement on diagnosis (AB yes/SI yes); agreement on no diagnosis (AB no/SI no); disagreement where AB is yes and SI is no; and disagreement where AB is no and SI is yes. **Notably, although differences were statistically significant due to the large sample size, the effect sizes (e.g., percentage differences observed in Appendix 4a) were not clinically meaningful.**

Abbreviations: MDD, major depressive disorder; GAD, generalised anxiety disorder; AB, algorithm-based diagnoses; SI, single-item

*p < 0.05, **p < 0.01, ***p < 0.001

##### Table 4b-2. Chi-squared results comparing differences by sex for binary agreement and disagreement results for each disorder.

| **disorder** | **statistic** | **df** | **p-value** |
| --- | --- | --- | --- |
| MDD | 3.687834 | 1 | 0.0548108 |
| Any anxiety | 1.754550 | 1 | 0.1853058 |
| GAD | 245.148533 | 1 | p < 2.2e-16*** |
| Specific phobia | 59.421239 | 1 | 1.27e-14*** |
| Social anxiety disorder | 10.073226 | 1 | 0.0015044** |
| Panic attacks | 213.092037 | 1 | p < 2.2e-16*** |
| Panic disorder | 24.016649 | 1 | 0.0000010*** |
| Agoraphobia | 16.372232 | 1 | 0.0000520*** |

Table 4b-2 displays the result of chi-squared analyses comparing the agreement and disagreement between algorithm-based (AB) and single-item (SI) diagnoses for males (N = 15,786) and females (N = 42,614) for each disorder. The outcome included 2 levels of agreement/disagreement: agreement (AB yes/SI yes or AB no/SI no); or disagreement (AB yes/SI no or AB no/SI yes). **Notably, although differences for the individual anxiety disorders were statistically significant due to the large sample size, the effect sizes (e.g., percentage differences observed in Appendix 4a) were not clinically meaningful.**

Abbreviations: MDD, major depressive disorder; GAD, generalised anxiety disorder; AB, algorithm-based diagnoses; SI, single-item

*p < 0.05, **p < 0.01, ***p < 0.001
